# Improving estimates of epidemiological quantities by combining reported cases with wastewater data: a statistical framework with applications to COVID-19 in Aotearoa New Zealand

**DOI:** 10.1101/2023.08.14.23294060

**Authors:** Leighton M. Watson, Michael J. Plank, Bridget A. Armstrong, Joanne R. Chapman, Joanne Hewitt, Helen Morris, Alvaro Orsi, Michael Bunce, Christl A. Donnelly, Nicholas Steyn

## Abstract

**Background:** Timely and informed public health responses to infectious diseases such as COVID-19 necessitate reliable information about infection dynamics. The case ascertainment rate (CAR), the proportion of infections that are reported as cases, is typically much less than one and varies with testing practices and behaviours, making reported cases unreliable as the sole source of data. The concentration of viral RNA in wastewater samples provides an alternate measure of infection prevalence that is not affected by clinical testing, healthcare-seeking behaviour or access to care.

**Methods:** We constructed a state-space model with observed data of levels of SARS-CoV-2 in wastewater and reported case incidence and estimated the hidden states of *R* and CAR using sequential Monte Carlo methods.

**Results:** Here, we analysed data from 1 January 2022 to 31 March 2023 from Aotearoa New Zealand. Our model estimates that *R* peaked at 2.76 (95% CrI 2.20, 3.83) around 18 February 2022 and the CAR peaked around 12 March 2022. We calculate that New Zealand’s second Omicron wave in July 2022 was similar in size to the first, despite fewer reported cases. We estimate that the CAR in the BA.5 Omicron wave in July 2022 was approximately 50% lower than in the BA.1/BA.2 Omicron wave in March 2022.

**Conclusions:** Estimating *R*, CAR, and cumulative number of infections provides useful information for planning public health responses and understanding the state of immunity in the population. This model is a useful disease surveillance tool, improving situational awareness of infectious disease dynamics in real-time.

**Plain Language Summary:** To make informed public health decisions about infectious diseases, it is important to understand the number of infections in the community. Reported cases, however, underestimate the number of infections and the degree of underestimation likely changes with time. Wastewater data provides an alternative data source that does not depend on testing practices. Here, we combined wastewater observations of SARS-CoV-2 with reported cases to estimate the reproduction number (how quickly infections are increasing or decreasing) and the case ascertainment rate (the fraction of infections reported as cases). We apply the model to Aotearoa New Zealand and demonstrate that the second wave of infections in July 2022 had approximately the same number of infections as the first wave in March 2022 despite reported cases being 50% lower.

## 1 Introduction

Understanding and predicting the trajectory of infectious diseases is important in planning an effective public health response. Reported case data depend heavily on testing modalities and practices which typically change over time, resulting in considerable uncertainty in the case ascertainment rate (CAR; the fraction of infections that are officially reported). During the COVID-19 pandemic, many countries relied primarily on symptom-based testing programmes to inform situational awareness and public health responses. In Aotearoa New Zealand, the CAR for COVID-19 has been influenced by factors such as access to testing, a shift from healthcare worker-administered polymerase chain reaction (PCR) tests to self-administered rapid antigen tests (RATs), reduction in rates of symptomatic and severe disease due to rising population immunity, relaxation of testing requirements and recommendations, and/or lack of perceived need to test or ‘pandemic fatigue’ [1–3]. As a result, over time, officially reported cases of COVID-19 have become a less reliable measure of levels of SARS-CoV-2 infection.

Data on hospital admissions and deaths are more consistent and are less affected by testing practices and behavioural change than reported cases but are subject to additional delays [4] that limit their usefulness for understanding disease dynamics. Infection prevalence surveys [5] that aim to regularly test a representative sample of the population are the gold-standard for tracking the spread of an infectious disease, but these surveys are resource intensive, making them harder to justify as countries move out of the acute phase of the pandemic. The UK was the only country to implement regular representative national SARS-CoV-2 prevalence surveys [6, 7] and there are no current plans for similar surveys in New Zealand.

Wastewater surveillance, where levels of SARS-CoV-2 RNA in wastewater samples are measured, can provide additional data on the prevalence of the virus that are unaffected by individual testing and self-reporting behaviours. Wastewater surveillance (also known as wastewater-based epidemiology or WBE) also has the potential to contribute to an integrated global network for disease surveillance [8–10]. These data, however, can be highly variable and subject to other biases, such as rainwater dilution, sampling methodologies, and changing locations of selected sampling sites. To realise this potential, appropriate models and analytical tools are needed to deliver epidemiological insights from raw data.

Two previous studies have presented novel methodology for the real-time estimation of the effective reproduction number using wastewater data [11, 12], while others have leveraged or extended these methods [13–16]. One study used reported cases to estimate the reproduction number and then fitted a model to estimate this quantity from wastewater data [17]. Another study used wastewater data to fit a mathematical model of multiple viral strains [18] from which estimates of the reproduction number can be derived. Other studies have analysed wastewater data but did not use it to estimate the reproduction number [19, 20]. Only [11] presented a model for simultaneously considering clinical and wastewater data, however they assume a fixed ascertainment rate. No previous work has combined wastewater-based epidemiology with reported cases to infer changes in case ascertainment over time.

Semi-mechanistic models based on the renewal equation are a popular method for epidemic forecasting and estimation of the instantaneous reproduction number [21–23]. Such methods are robust to constant under-ascertainment of cases, but may be biased by rapid changes in CAR and cannot provide any information about the total number of infections. In this paper, we extend the renewal equation framework [21–23] for reproduction number estimation to incorporate wastewater time-series data. The model treats the instantaneous reproduction number and CAR as hidden states and reported cases and quantity of viral RNA in wastewater as observed states. We use a sequential Monte Carlo approach to infer the hidden states. We apply the model to national data from Aotearoa New Zealand on reported COVID-19 cases and the average number of SARS-CoV-2 genome copies per person per day measured in municipal wastewater samples between January 2022 and March 2023. Because the relationship between infections and wastewater concentration is only determined in the model up to an overall scaling constant, it cannot be used to infer the absolute CAR but can be used to estimate relative changes in case ascertainment over time. The model is designed to be regularly updated as new data become available, producing real-time estimates of the effective reproduction number and relative change in CAR. The model has been used to support situational awareness via regular reports to the New Zealand Ministry of Health from November 2022 to date.

From March 2020 until December 2021 New Zealand used strict border controls and intermittent non-pharmaceutical interventions to suppress and eliminate transmission of SARS-CoV-2. By the beginning of 2022, there had been a cumulative total of around 3 confirmed cases of COVID-19 per 1,000 people and around 90% of the population over 12 years old had received at least two doses of the Pfizer-BioNTech vaccine. From October 2021, interventions were progressively eased and in January 2022 the B.1.1.529 (Omicron) variant began to spread in the community, causing the first large wave of infection. Since then community transmission has been sustained, with multiple further waves of infection being driven by various Omicron subvariants. Between 1 January 2022 and 31 March 2023, there was a cumulative total of around 440 confirmed cases per 1,000 people, most of which were from self-administered RATs. During this period, SARS-CoV-2 concentration was regularly measured at various wastewater treatment plants, providing an additional data source on changes in community prevalence over time.

We model the epidemic dynamics and the observed case and wastewater data at the national level, aggregating over New Zealand’s population of 5.1 million and ignoring regional variations. This is similar to other studies that have aggregated regional case and/or wastewater data to produce national-level estimates in countries with a comparable population size [24–26]. Our methodology could, in principle, be applied at a finer geographical scale, although this would come at the cost of higher levels of noise.

## 2 Materials and Methods

### 2.1 Data

National daily reported cases of COVID-19 were obtained from the New Zealand Ministry of Health [27]. Until February 2022, these cases were diagnosed solely by healthcare-administered PCR testing. From February 2022, in response to the rapid increase in reported cases, RATs were widely distributed. Since then, the vast majority of reported cases have been from self-administered RATs, with results reported via an online portal. Hence, data on the number of tests conducted are not available. Reported cases are shown in Figure 1. As these data exhibit a clear day-of-the-week effect we remove the weekly trend before fitting the model (see Supplementary Material section 1.2 for details).

**Figure 1.**
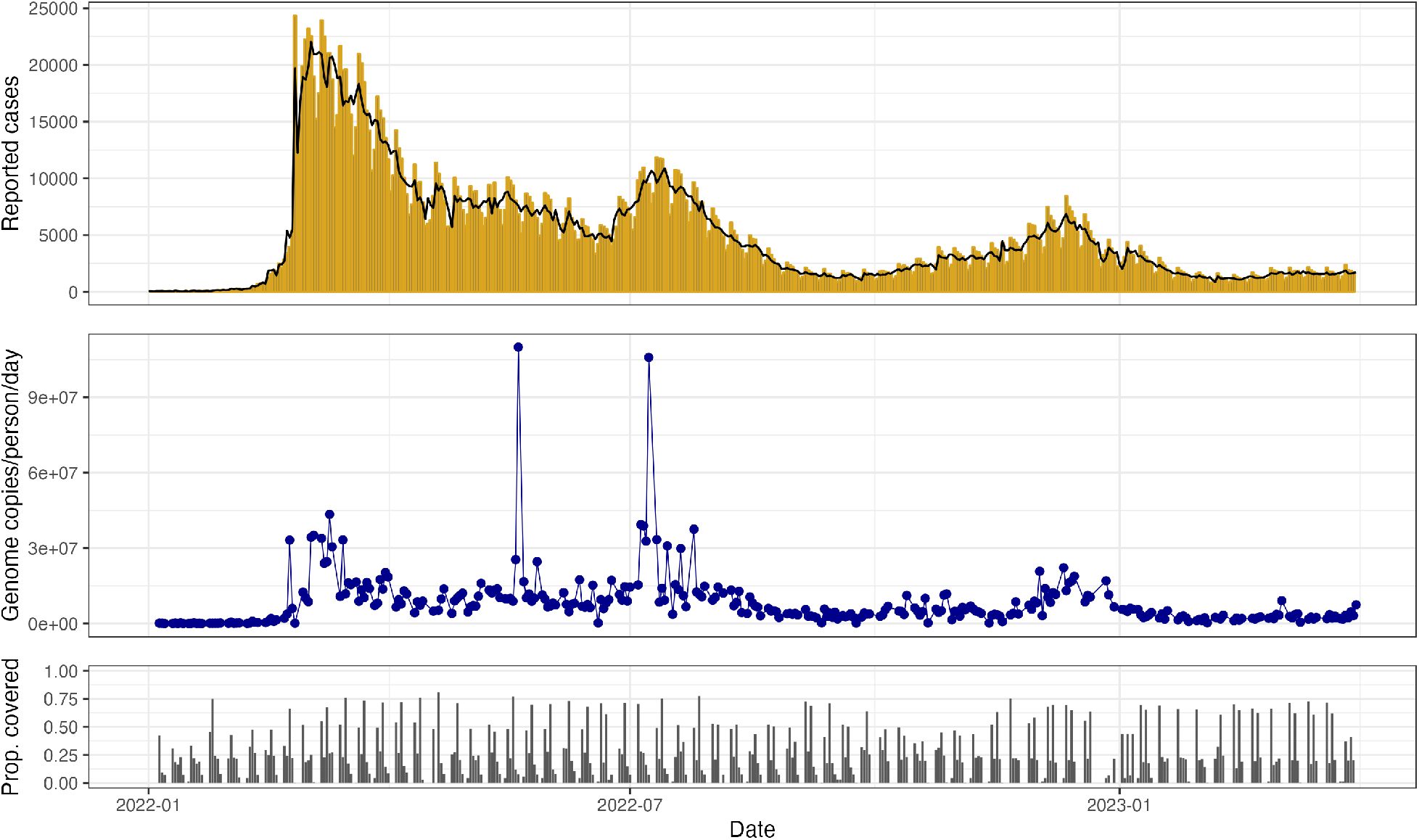
Reported daily cases of COVID-19 (upper), SARS-CoV-2 genome copies per person per day in sampled wastewater (middle), and proportion of the total population covered by sampled wastewater catchments (lower), between 1 January 2022 and 31 March 2023 in Aotearoa New Zealand. The black line in the upper plot shows the adjusted case series with the multiplicative day-of-the-week effect removed (see Supplementary Material section 1.2). The two outliers in wastewater data arise from estimates of a high wastewater flow-rate in Wellington following high rainfall. Since rainfall is a source of noise in wastewater sampling we retain these samples in our analysis. Reported case data were obtained from the New Zealand Ministry of Health [27] and wastewater data were obtained from ESR [28].

SARS-CoV-2 concentration data from wastewater samples tested by the Institute for Environmental Science and Research (ESR) were used for this study [28]. Wastewater samples were collected every week at municipal wastewater treatment plants located throughout the country, serving communities with populations ranging from 400 to over 500,000 people. Typically 70-90% of the national population connected to reticulated wastewater was covered by wastewater sampling in any given week (60-124 sites, usually sampled twice per week). Each site-level measurement was normalised to provide an estimate of the number of genome copies per person per day for that site (see Supplementary Material section 1.1). Typically multiple sites were sampled per day and, for each day that had at least one sample, we calculated the catchment-population-weighted average of the genome copies per person (see Figure 1). Because we do not attempt to model regional variations, we assumed this provided a series of representative observations of the average concentration of genomic material in the national wastewater (see also Section 2.2).

### 2.2 Hidden state model

We construct a state-space model (Figure 2) consisting of time-varying hidden states (the instantaneous reproduction number *R*_*t*_, daily case ascertainment rate *CAR*_*t*_, and daily infection incidence *I*_*t*_) and time-varying observed states (daily reported cases of COVID-19 *C*_*t*_ and daily wastewater observations *W*_*t*_). We use subscript *s* : *t* to refer to all values between day *s* and *t* inclusive.

**Figure 2.**
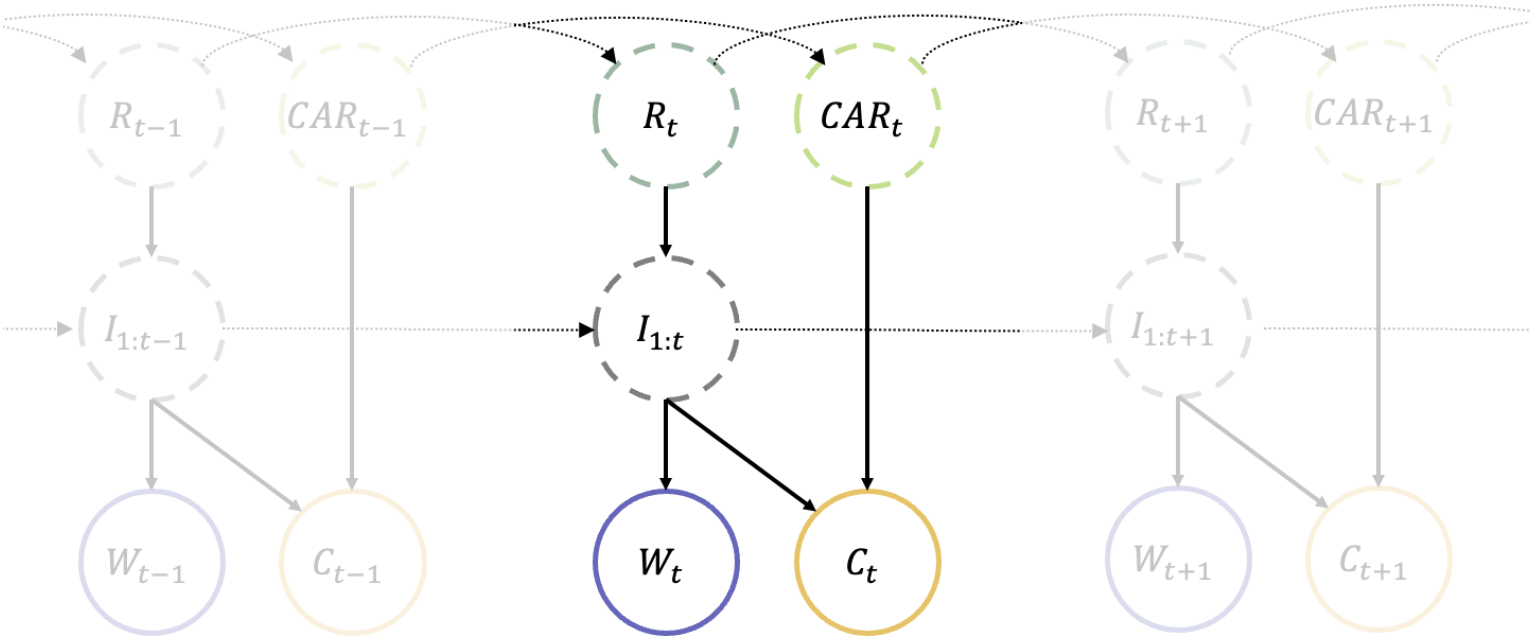
Diagram of the state-space model showing the dependency between hidden-states (dashed circles) and the observed data (solid circles). *R*_*t*_ is the instantaneous reproduction number on day *t, CAR*_*t*_ is the case ascertainment rate on day *t, I*_*t*_ is the number of new infections on day *t, C*_*t*_ is the number of reported cases on day *t*, and *W*_*t*_ is the observed wastewater, measured as the total genome copies per person per day for the sites that were sampled on day *t. I*_1:*t*_ denotes the set of states *{I*_1_, *I*_2_, …, *I*_*t*_*}*. In practice the current infections *I*_*t*_, reported cases *C*_*t*_ and wastewater *W*_*t*_ depend only on recent values of *I*_*t*_ as specified by the generation interval distribution, the infection-to-reporting distribution, and infection-to-shedding distribution respectively (see Methods).

We assume the hidden states *R*_*t*_ and *CAR*_*t*_ follow independent Gaussian random walks, encoding the fact we expect them to vary continuously over time. We also assume that the hidden state *I*_*t*_ follows a Poisson renewal process, a simple epidemic model commonly used when estimating *R*_*t*_ [21]. Thus our state-space transitions are governed by:

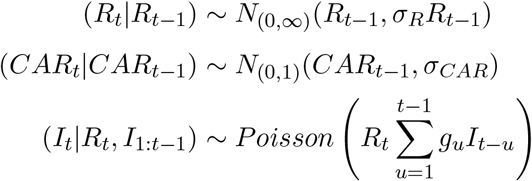

Parameters *σ*_*R*_ and *σ*_*CAR*_ determine how quickly *R*_*t*_ and *CAR*_*t*_ vary. The standard deviation of the transition distribution for *R*_*t*_ → *R*_*t*+1_ is given by *σ*_*R*_*R*_*t*_, which means that *R*_*t*_ varies more rapidly at larger values. The distribution for *R*_*t*_ was truncated on (0, ∞) and for *CAR*_*t*_ on (0, 1). Finally, *g*_*u*_ is the pre-determined generation time distribution, describing the proportion of transmission events that occur *u* days after infection (see Supplementary Material section 2.7).

We assume that the expected number of reported cases 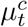 at time *t* is equal to *CAR*_*t*_ multiplied by the convolution of past infections with the infection-to-reporting distribution *L*_*u*_:

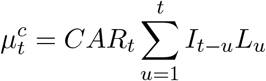

Similarly, we assume that the expected number of genome copies 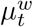 detected per person at time *t* is equal to the convolution of past infections with the infection-to-shedding distribution *ω*_*u*_, multiplied by a fixed parameter *a* representing the average total detectable genome copies shed into the wastewater by an infectious individual, divided by total national population size *N* :

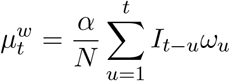

We model reported cases using a negative binomial distribution:

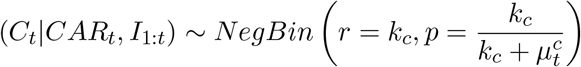

which has mean 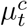 and variance 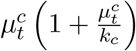. A negative binomial distribution is used to account for noise in the observations beyond that predicted by a binomial distribution. This is a common choice in other methods of reproduction number estimation [23, 29].

The observed wastewater data *W*_*t*_ is the total genome copies per person from the wastewater sites sampled on day *t*. We model this using a shape-scale gamma distribution:

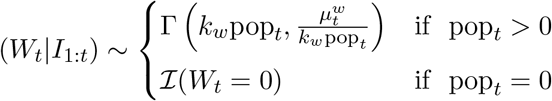

which has mean 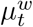 and variance 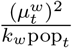. This assumes that the observed daily data are independent draws from the national distribution, which may not hold if there are regional differences between the subsets of sites that are sampled on different days. In practice, any such differences will be absorbed into the variance of the daily observation distribution via fitting of the dispersion parameter *k*_*w*_. Since we marginalise out the effect of this parameter when presenting results, the increased uncertainty associated with regional variability is propagated through to the credible intervals. The variable pop_*t*_ refers to the total population in the catchment areas of the sampled wastewater sites on day *t*. Setting the variance of the observation distribution to be inversely proportional to pop_*t*_ allows the model to account for increased variability around the national mean on days when fewer or smaller sites were sampled. ℐ is the indicator function, so on days when no sites were sampled, the probability of observing no wastewater samples is set to 1, and the model fits to case data alone.

Consistent with previous models [30, 31], this formulation assumes that the expected population shedding rate is proportional to the number of infected individuals, with observations drawn from a distribution around this mean. We used a gamma distribution, which is a reasonably flexible choice for a non-negative continuous random variable. However other distributions could be considered, such as a Weibull or log-normal.

In the absence of additional information we are unable to estimate *α*, which is proportional to the average total genome copies shed by an infected individual over the course of their infection. This means we are unable to estimate the absolute value of *CAR*_*t*_. Instead, we run the model with a range of different values for *α*, and estimate the change in *CAR*_*t*_ relative to its initial value. This additionally requires the assumption that *α* is constant over time, which is unlikely to be true in general and is a key limitation of our model (see Discussion).

In practice, the range of values of *α* that we used (see Table 1) was chosen by calibrating model output for the number of infections with external sources of information. Firstly, we compared model output to the number of cases in a cohort of around 20,000 border workers who were tested weekly between January and July 2022 [36]. Secondly, around 40% of all 20-25-year-olds (an age group unlikely to have a higher CAR than older adults) reported a case of COVID-19 in the 6 months from 1 February to 31 July 2022 [27]. This suggests that the overall CAR for this period was likely to be at least 0.4, which translates to an approximate upper bound of 4 million for the total cumulative number of infections up to 31 July 2022. Neither of these observations definitively determines the number of infections as they are subject to approximation, bias and uncertainty, but they nevertheless serve to bracket the likely range of values for the parameter *α*.

**Table 1:** Parameter values used in the model. The infection-to-reporting and infection-to-shedding distributions are calculated as convolutions of the incubation period distribution [32] and the onset-to-reporting and onset-to-shedding distribution [33] respectively (see Supplementary Material section 2.7).

| Parameter | Symbol | Value |
| --- | --- | --- |
| Coefficient of variation of $R_t$ transitions | $\sigma_R$ | Fitted |
| Std dev. of $CAR_t$ transitions | $\sigma_{CAR}$ | Fitted |
| Reported cases tuning parameter | $k_c$ | Fitted |
| Wastewater tuning parameter | $k_w$ | Fitted |
| Generation time distribution [34, 35] | $g_u$ | Mean = 3.3 days, s.d. = 1.3 days |
| Infection-to-reporting distribution | $L_u$ | Mean = 5.8 days, s.d. = 2.6 days |
| Infection-to-shedding distribution | $\omega_u$ | Mean = 5.2 days, s.d. = 2.9 days |
| Average total genome copies per infection | $\alpha$ | $3 \times 10^9$ [$2 \times 10^9$, $4 \times 10^9$] |
| Fixed-lag resampling window | $h$ | 30 days |

The infection-to-reporting and infection-to-shedding distributions are calculated as the convolution of the incubation period distribution with the onset-to-reporting and onset-to-shedding distribution respectively. The incubation period is modelled as a Weibull distribution with mean 2.9 days and standard deviation 2.0 days [32]. The onset-to-reporting distribution is estimated empirically from New Zealand case data extracted on 16 September 2022, representing over 1.2 million cases, and has mean 1.8 days and standard deviation 1.8 days. The onset-to-shedding distribution comes from [33] and has mean 0.7 days and standard deviation 2.6 days. The resulting infection-to-reporting distribution has mean 5.8 days and standard deviation 2.6, and the resulting infection-to-shedding distribution has mean 5.2 days and standard deviation 2.9 days (see Supplementary Figure S1).

The model is solved using a bootstrap filter [37] with fixed-lag resampling. This produces estimates for the marginal posterior distribution of the hidden states at each time step. The random walk step variance parameters (*σ*_*R*_ and *σ*_*CAR*_) and observation variance parameters (*k*_*c*_ and *k*_*w*_) are estimated using a particle marginal Metropolis Hastings Markov chain Monte Carlo method. We use uninformative uniform prior distributions for these parameters, with the exception of *σ*_*CAR*_, where we use an informative prior distribution to ensure an appropriate level of smoothness in our estimates of *CAR*_*t*_. Different parameter values are fitted in three-month blocks to allow for some variation over time. See Supplementary Material section 2 for further details of the numerical method. Code and data to reproduce the results are provided at https://github.com/nicsteyn2/NZWastewaterModelling.

## 3 Results

### Reproduction number, relative case ascertainment, and infection incidence

The estimated value of the reproduction number *R*_*t*_ (Figure 3a) increased from around 1 at the beginning of 2022 to a peak of 2.46 (95% CrI 2.04, 3.20) on 18 February 2022 (95% CrI 10 Feb, 23 Feb), corresponding to the sharp increase in cases seen during the first Omicron wave, which was a mixture of the BA.1 and BA.2 variants [38]. The estimated value of *R*_*t*_ dropped below 1 on 1 March 2022 (95% CrI 25 Feb, 5 Mar) and infection incidence peaked on 28 February 2022 (95% CrI 23 Feb, 7 Mar), suggesting this is when the wave peaked.

**Figure 3.**
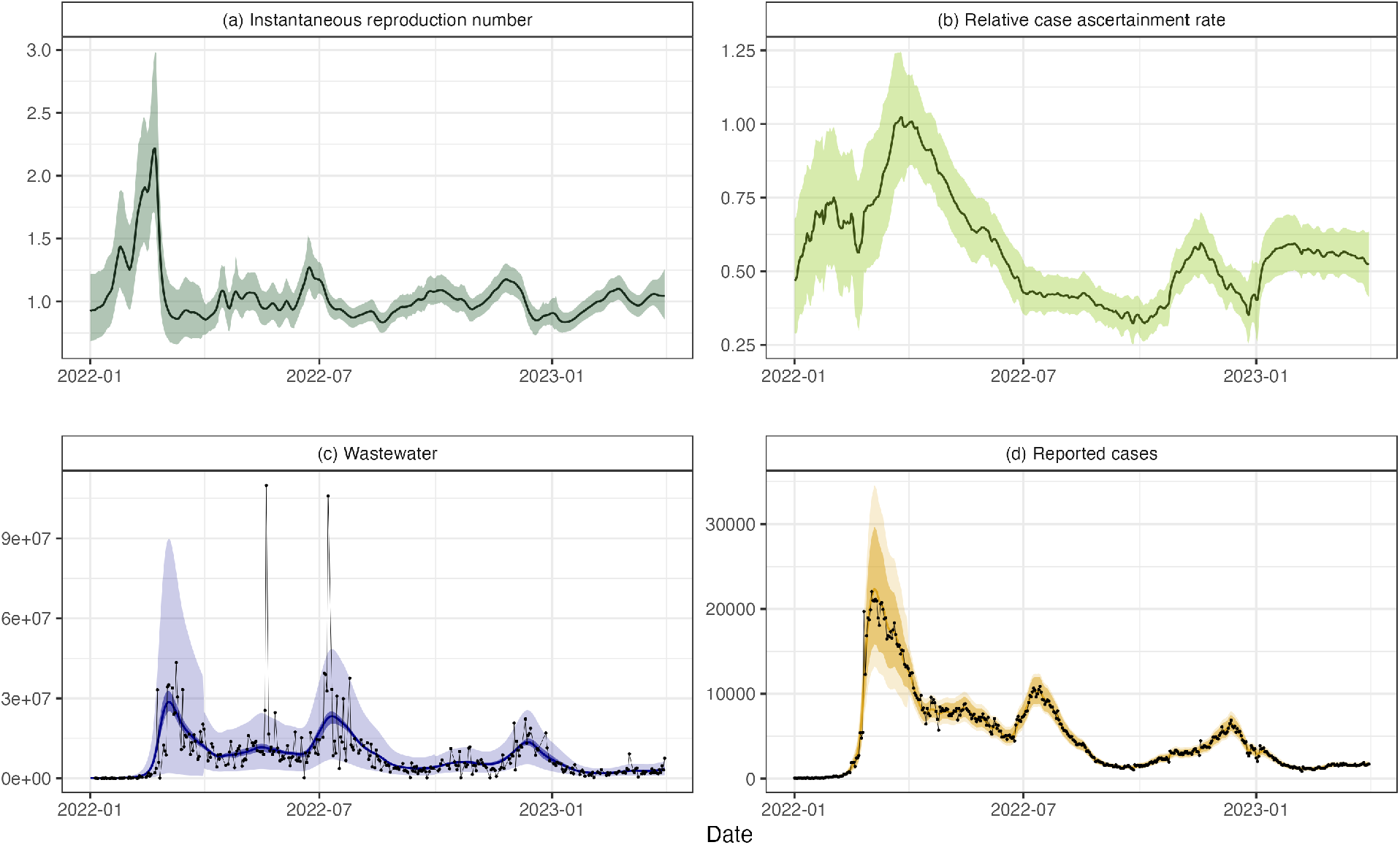
Results for New Zealand data from 1 January 2022 to 31 March 2023. (a) instantaneous reproduction number *R*_*t*_, (b) relative case ascertainment rate (compared to the central estimate on 1 April 2022), (c) wastewater data *W*_*t*_ measured in genome copies per person per day and (d) reported cases *C*_*t*_. Results assume the average total shedding per infection does not vary over time (*α* = 3 *×* 10^9^). Solid lines present central estimates. Shaded regions show 95% credible intervals on the value of the hidden states (subplots a and b), and 95% credible intervals on the expected reported cases and wastewater data (darker shaded regions in subplots c and d) and 95% credible intervals on the prediction distribution for wastewater data and reported cases (lighter shaded regions in subplots c and d). Black dots show the observed data.

The estimated CAR (Figure 3b) increased rapidly between mid-February and mid-March 2022. RATs became widely available for the first time in the last week of February 2022. This likely led to a significant increase in case ascertainment as the testing system, which had previously relied solely on laboratory-processed PCR tests, had become overwhelmed [3]. The estimated CAR approximately halved between April and July 2022, when a second wave of infection caused by the BA.5 Omicron subvariant [36, 38] occurred. This second wave was visible in both reported cases and wastewater sampling, with estimated peak infections occurring on 7 July 2022 (95% CrI 3 Jul, 12 Jul). The estimated CAR increased somewhat between mid 2022 and early 2023, with a noticeable dip in December 2022, possibly reflecting reduced testing during the Christmas and summer school holiday period (from mid-December to late-January/early-February). Alternatively, the estimated increase in CAR from mid-2022 could be explained by a decrease in the average genome copies shed by an infected individual *α*, although without further information we are unable to discern changes in *α*. Overall, the model provided a reasonably good fit to the observed data on cases and wastewater (Figure 3c-d).

Figure 4a-b shows the estimated daily incidence and cumulative infections for three values of *α*, corresponding to estimated CAR values on 1 April 2022 of 0.42 (95% CrI 0.35, 0.50), 0.61 (95% CrI. 0.51, 0.71), and 0.80 (95% CrI. 0.67, 0.93), for *α* = 2 *×* 10^9^, 3 *×* 10^9^, and 4 *×* 10^9^ respectively. For comparison, the graphs also show the number of cases per capita in a cohort of approximately 20,000 border workers who were tested weekly between January and July 2022 [36], scaled according to population size. These data were used to help inform the range of values of *α* selected (see Methods).

**Figure 4.**
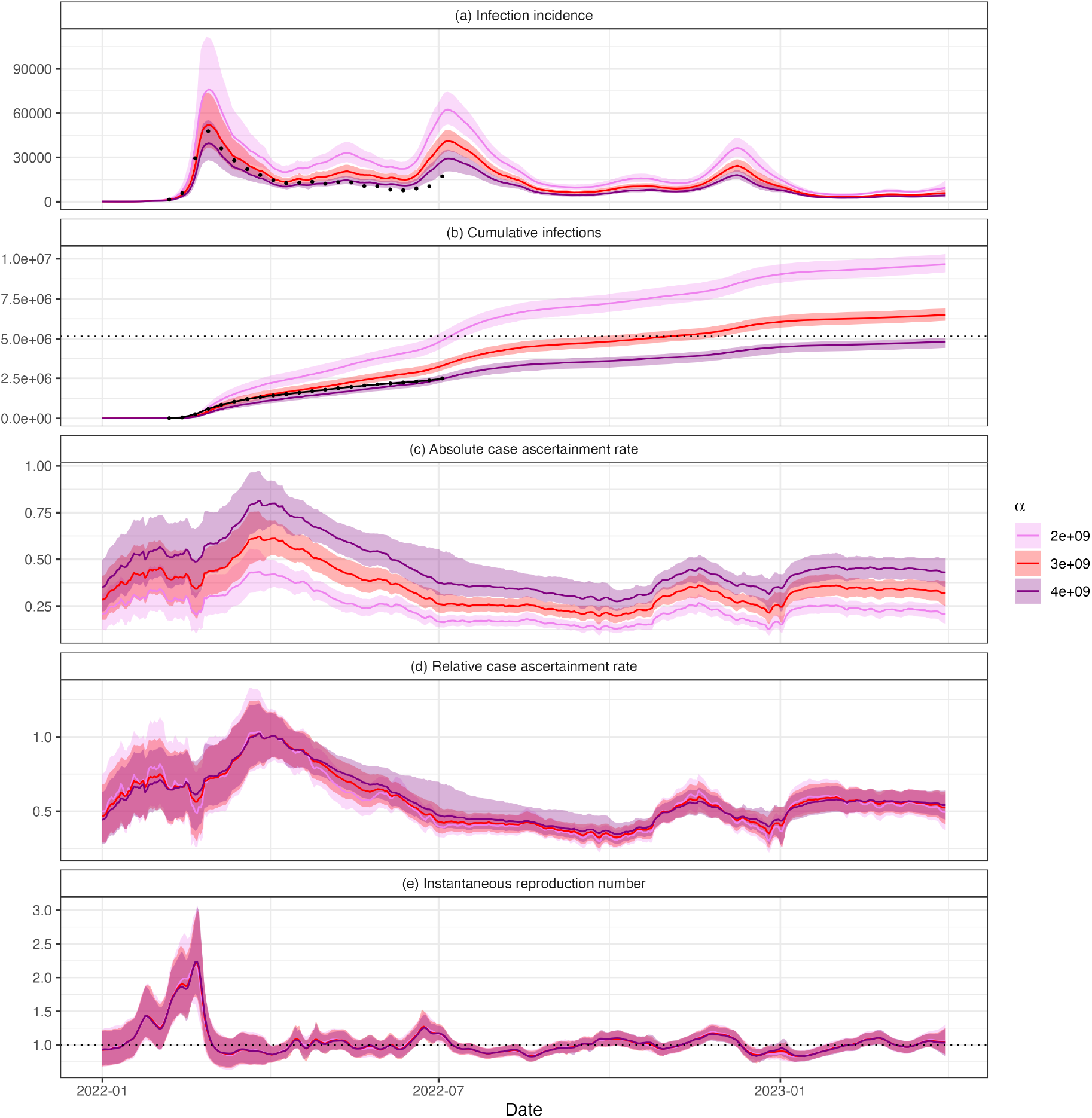
Estimated (a) daily infections *I*_*t*_, (b) cumulative infections 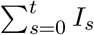, (c) case ascertainment rate *CAR*_*t*_, (d) relative case ascertainment rate (compared to the central estimate on 1 April 2022), and (e) instantaneous reproduction number, *R*_*t*_. Results are presented for three values of *α*: 2 *×* 10^9^, 3 *×* 10^9^, and 4 *×* 10^9^. Solid lines show central estimates and coloured regions are the 95% CrIs. Estimates and credible intervals on cumulative infections are calculated by taking cumulative sums of the estimates and credible intervals in panel (a). Black dots in panels (a) and (b) show the number of per capita cases in a cohort of regularly-tested border workers, scaled according to population size. The horizontal dashed black lie in panel (b) shows the New Zealand population at the end of 2022 (5.15 million people) [39]. While changing *α* results in different estimates of infections and absolute CAR, the relative CAR and reproduction number estimates are robust to different values, provided *α* remains relatively constant.

Whilst peak reported cases (adjusted for the day-of-the-week effect) in the second wave were only 49% of the peak in the first wave (10,879 vs 22,038 respectively), under the assumption of constant *α*, the central estimate from the model suggests that true infections peaked at approximately 78% of the peak of the initial wave (Figure 4a). Figure 4c-e shows the estimated absolute and relative CAR and *R*. These panels show that, while we are uncertain about the absolute level of infections and CAR, the relative CAR and reproduction number estimates are robust to reasonable choices for (constant) *α*.

Fitting the model to case data alone instead of cases and wastewater (see Supplementary Figure S7) produced qualitatively similar estimates of *R*_*t*_, but with greater temporal fluctuations. Fitting the model to wastewater data alone led to substantially wider credible intervals, although the overall trend was similar. Estimates of the relative CAR are only possible when fitting to case and wastewater data simultaneously.

### Parameter estimates

The estimated standard deviation *σ*_*R*_ of the random walk on *R*_*t*_ was greatest in the first time period (1 Jan – 31 Mar 2022) – see Table 2. This is unsurprising as it coincided with the rapid increase and then decrease in incidence associated with the first Omicron wave. *σ*_*R*_ decreased in the second period (1 Apr – 30 Jun 2022) and then remained relatively constant throughout the remaining periods (1 Jul 2022 – 31 Mar 2023). The estimated standard deviation *σ*_*CAR*_ of the random walk on *CAR*_*t*_ was also estimated to be greatest in the first time period, although this is primarily because we applied a prior distribution with a higher mean in this period (see Supplementary Material section 2.5).

**Table 2:**
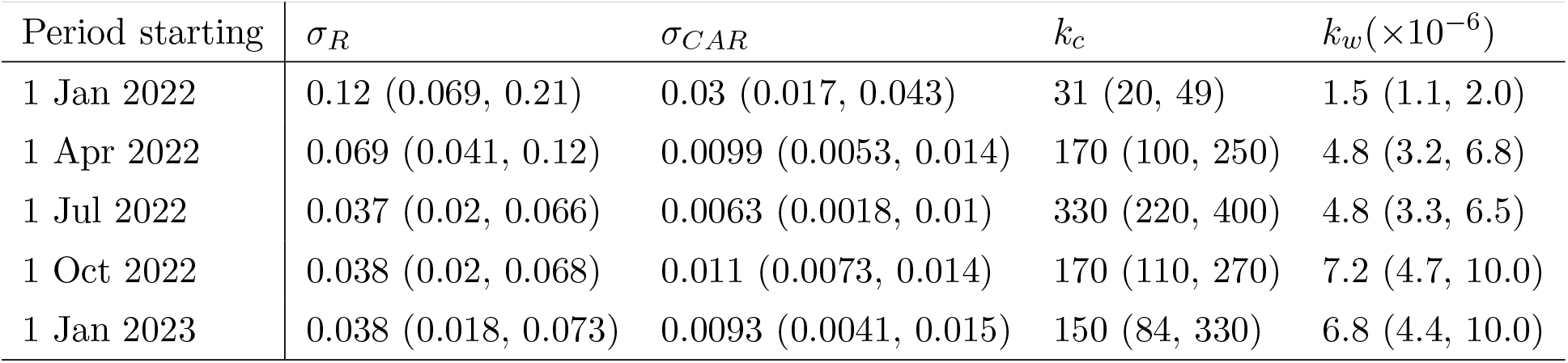
Central estimates and 95% CrIs for estimated model parameters in each time period. Dates in the ‘Period’ column are the start date for the three-month period. All outputs presented to 2 s.f. Higher values of *σ*_*R*_ and *σ*_*CAR*_ suggest *R*_*t*_ and *CAR*_*t*_ vary faster. Higher values of *k*_*c*_ and *k*_*w*_ indicate a lower variance in the corresponding observation distribution. Note a different prior distribution was used for *σ*_*CAR*_ in the first period (see Supplementary Material, section 2.5), which may also impact estimates of other parameters in this period.

| Period starting | $\sigma_R$ | $\sigma_{CAR}$ | $k_c$ | $k_w (\times 10^{-6})$ |
| --- | --- | --- | --- | --- |
| 1 Jan 2022 | 0.12 (0.069, 0.21) | 0.03 (0.017, 0.043) | 31 (20, 49) | 1.5 (1.1, 2.0) |
| 1 Apr 2022 | 0.069 (0.041, 0.12) | 0.0099 (0.0053, 0.014) | 170 (100, 250) | 4.8 (3.2, 6.8) |
| 1 Jul 2022 | 0.037 (0.02, 0.066) | 0.0063 (0.0018, 0.01) | 330 (220, 400) | 4.8 (3.3, 6.5) |
| 1 Oct 2022 | 0.038 (0.02, 0.068) | 0.011 (0.0073, 0.014) | 170 (110, 270) | 7.2 (4.7, 10.0) |
| 1 Jan 2023 | 0.038 (0.018, 0.073) | 0.0093 (0.0041, 0.015) | 150 (84, 330) | 6.8 (4.4, 10.0) |

The estimated variance parameters, *k*_*c*_ and *k*_*w*_, for cases and wastewater observations, were lowest in the first time period (1 Jan 2022 – 31 Mar 2022). This implies there is more variability in the data that is not explained by the model in this time period, possibly as a consequence of the sharper variations in incidence compared to the later time periods. A less consistent weekly pattern in reported cases during the first time period, and higher levels of noise in wastewater observations at the low concentrations seen at the beginning of 2022, could also be contributing factors.

## 4 Discussion

Wastewater-based epidemiology has been used globally for COVID-19 surveillance and has been shown to be a useful public health tool for policy and public health responses [40]. We have presented a semi-mechanistic model that combines reported cases with wastewater data to estimate the time-varying reproduction number and CAR. This work demonstrates the value of wastewater-based epidemiology and how the additional data that it provides can be combined with traditional monitoring (e.g., reported cases) to learn more about the state of an epidemic, disease dynamics, and the true number of infections in the community. This provides useful information to inform the public health response.

To make reliable estimates of the state of the epidemic from reported cases, it is essential to understand how case ascertainment changes with time. For example, are there fewer cases because there are fewer infections or because fewer people are reporting? We applied our model to national data from Aotearoa New Zealand and derived insights into changes in case ascertainment that would not be possible using case data alone. Reported cases during the second wave in July 2022 were significantly lower than in the first wave in February and March 2022. However, the model inferred that there was a substantial drop in case ascertainment between these waves, and the true number of infections was likely more similar in each wave. The reduced CAR during the second and subsequent waves may have been due to a higher number of reinfections with individuals displaying fewer symptoms or due to “pandemic fatigue” and reduced compliance with public health measures, including testing. This type of insight would not be possible without regular wastewater surveillance data and without a robust analytical framework in which to integrate these data with traditional epidemiological data streams.

We applied our model to the first period of widespread community transmission of SARS-CoV-2 in New Zealand. During this time, rapid antigen tests were freely available to everyone, there was a requirement to report positive results, and a mandatory isolation period for cases with financial support via employers. Partly as a result of these factors, the CAR, while lower than in the previous elimination phase, was still reasonably high. The mandatory isolation period was removed in September 2023, which led to a substantial drop in case ascertainment. For the datasets we considered, similar (albeit noisier) estimates for the reproduction number could be obtained from case data alone. However, in a context where case ascertainment is low and/or unrepresentative, wastewater data are likely to add even greater value compared to using reported cases. In contrast, in a low-prevalence context (e.g. pre-Omicron in New Zealand), applicability of the method would be constrained by the amount of noise in the wastewater data. In this situation, wastewater surveillance may better used for presence/absence monitoring, for example as an early warning system for the presence of infection in specific catchments, as opposed to quantitative estimation [41].

Strengths of our model include the fact that it has relatively minimal data requirements, requiring only time series for reported cases and wastewater concentrations. The model can be fitted to datasets in which different sites are sampled on different days and some days have no observed data. This means that it could be readily applied in other jurisdictions with wastewater surveillance programs, either for SARS-CoV-2 or other pathogens such as influenza viruses [40, 42]. It is a relatively simple model with minimal mechanistic assumptions and parsimonious parameterisation. This avoids the need for assumptions about time-varying contact patterns, transmission rates, and the level of prior immunity that are required by more complex mechanistic models. The model presented here was operationalised by ESR in late 2022 and results for *R*_*t*_ and relative CAR are regularly provided to the Ministry of Health to inform situational awareness and decision-making.

There are several limitations to this model and the results. We assumed that the average number of genome copies shed by an infected individual (represented the parameter *α*) was constant between January 2022 and March 2023 and did not depend on the infecting variant or history of prior infection or vaccination. It is possible that some of the inferred changes in CAR may be partly explained by these factors. For example, some of the inferred increase in case ascertainment between October and December 2022 may have been due to decreasing *α*, caused by a combination of new immune evasive subvariants displacing the previously dominant BA.5 variant [43] and/or an increase in the proportion of reinfections or asymptomatic infections [27]. Although estimates of viral shedding rates per infected individual are available [30, 31], the value of *α* may also depend on physical characteristics of the wastewater collection system, sample collection method, and the method used to quantify concentration of SARS-CoV-2 RNA in samples. Therefore, *α* is likely to vary between jurisdictions and will require recalibration using local data.

As we are unable to estimate the true value of *α*, we are unable to estimate the absolute CAR. Nonetheless, relative CAR is a useful metric and, given an estimated range of values for *α*, we are able to provide plausible bounds on the total number of infections (Figure 4).

Wastewater surveillance does not provide any information on how infections are distributed among population groups (e.g. age groups, ethnicity) and biases in self-administered testing mean that case counts are not representative either. This information is important for assessing the clinical burden of disease and addressing health inequities [44]. Thus, other approaches are needed to determine the distribution of disease burden, such as representative sampling [7, 45], cohort studies [46] or sentinel surveillance [47, 48]. Although wastewater surveillance could, in principle, be used to investigate differences in prevalence and case ascertainment between sites and/or regions, this would require adaptions to our method that are beyond the scope of this study.

As our model is flexible, future work could integrate hospitalisations (such as in [49]) and deaths data. In principle, this could allow the effects of varying CAR and varying rate of shedding per infection to be separated. However, this would additionally require the effects of age, immunity, ethnicity, and other variables on clinical severity to be accounted for.

Although national-level approaches to situational awareness and reproduction number estimation are common [25, 50, 51], particularly in countries such as New Zealand with a relatively small population size, this ignores regional variations. Results should therefore be interpreted as national averages, which could mask demographic and spatial heterogeneity. Our model could be implemented at a regional level so that local epidemic dynamics can be compared, although this would be subject to increasing levels of noise in the wastewater data at finer spatial scales. This paper has focused on modelling for inference: understanding epidemic dynamics that have already occurred. However, the state-space transition model coupled with the estimated parameters provides a natural method for forecasting [23, 52]. Forecasts generated using this state-space transition model naturally incorporate increasing uncertainty about the future reproduction number and CAR.

While this model has focused on COVID-19, there is a wealth of genetic information within municipal wastewater that could also benefit from modelling. The detection and concentration of viral, bacterial and anti-microbial resistance genes within wastewater have the ability to inform public health decision-making in a number of ways, especially as methodology is refined allowing more rapid turnaround times. As many jurisdictions seek to retain the wastewater capabilities they built during the pandemic phase of COVID-19 (and to diversify microbial targets), there is an ‘opportunity springboard’ to build tools that can predict the trajectories and spread of pathogens. Modelling has a key role to play in this journey.

## Data availability

Daily reported case data for Aotearoa New Zealand are available from the Ministry of Health at https://github.com/minhealthnz/nz-covid-data and seven-day average wastewater data are available from ESR at https://github.com/ESR-NZ/covid_in_wastewater.

Code to run the model and reproduce the results in this paper are available at https://github.com/nicsteyn2/NZWastewaterModelling.

## Acknowledgements

The authors acknowledge the role of the New Zealand Ministry of Health in supplying data in support of this work. The authors thank the wastewater treatment plant staff members who collected the wastewater samples and the ESR laboratory staff who processed and tested the samples used in this study. This work was funded by the New Zealand Ministry of Health and the Department of Prime Minister and Cabinet (DPMC). This work was supported by the NIHR HPRU in Emerging and Zoonotic Infections, a partnership between PHE, University of Oxford, University of Liverpool, and Liverpool School of Tropical Medicine (grant number NIHR200907 supporting C.A.D.). L. M. W. was supported by a Rutherford Foundation Postdoctoral Fellowship from New Zealand government funding, administered by the Royal Society Te Apārangi. N.S. acknowledges support from the Oxford-Radcliffe Scholarship from University College, Oxford, and the Engineering and Physical Sciences Research Council (EPSRC) Centre for Doctoral Training (CDT) in Modern Statistics and Statistical Machine Learning (Imperial College London and University of Oxford). We thank A. Maslov for supporting this research through studentship support for N.S.

