## Supporting Information for "Improving estimates of epidemiological quantities by combining reported cases with wastewater data: a statistical framework with applications to COVID-19 in Aotearoa New Zealand"

**4 Particle Marginal Metropolis-Hastings Outputs** **18**

### 1 Data

#### 1.1 Wastewater Sampling

Composite samples were collected by an autosampler, which collects a small volume of waste water at regular intervals over the course of a 24-hour period. When composite samples were not available, 'grab' samples were collected and ranged from a sample being taken at a single point in time to three samples taken over 30 minutes. Grab samples represent only the composition of the source at the time of collection and may not be as representative as a 24-hour composite sample collected by an autosampler. Following collection, samples were couriered overnight to the Institute of Environmental Science and Research (ESR) for processing.

Virus concentration was performed by SARS-CoV-2 detection and quantitation by RT-qPCR of the N-gene as described in [1]. Wastewater (250 mL) was concentrated to 1.25 mL, of which 0.2 mL was used for nucleic extraction. Six RT-qPCR replicates were performed for each sample. SARS-CoV-2 RNA was considered detected if any of the RT-qPCR replicates were positive. A result of 'not detected' meant that SARS-CoV-2 RNA was either absent from the sample or at a level too low to be reliably reported.

RT-qPCR data (quantification cycle values) were converted to genome copies per reaction using a standard curve and then to genome copies per litre of wastewater. Each sample is multiplied by the estimated volume of wastewater entering the wastewater treatment plant to estimate the total genome copies per day. These are summed over all sites sampled on that day and divided by the total volume of wastewater entering all sampled wastewater treatment plants to give daily estimates of genome copies per litre of wastewater. We also calculate the daily total population across sampled catchment sites.

The data and comprehensive details about collection and processing are available at `https://github.com/ESR-NZ/covid_in_wastewater` [2].

#### 1.2 Reported Cases

National daily reported cases of COVID-19 were obtained from the New Zealand Ministry of Health and are avaliable at `https://github.com/minhealthnz/nz-covid-data` [3]. Reported case data exhibit a clear day-of-the-week effect, which we remove in pre-processing using a simple linear regression model. The log-transformed daily case count was regressed against the day of the week and the data were then divided by the exponential of the regression coefficient for each day of the week. Adjusted daily case counts were then scaled so the total case count remains consistent. We perform this day-of-the-week adjustment in the following consecutive time windows: before 1 January 2022, 1 January to 28 February 2022, 1 March to 30 June 2022, 1 July to 30 September 2022, 1 October to 31 December 2022, and 1 January 2023 to 31 March 2023. These are mostly three months in duration, except that the boundary between the second and third windows was selected to coincide with the change from PCR-only testing to widespread availability and use of RATs [4], as this impacted the weekly reporting pattern.

Algorithmically, this seasonal adjustment is performed when the data are loaded. As it results in non-integer daily case counts, we round the outputs to the nearest integer. The functions to do this are included in the "loadNZData.jl" function at `https://github.com/nicsteyn2/`

`NZWastewaterModelling/tree/main/src`.

### 2 Model Derivation and Algorithms

#### 2.1 Fixed-Lag Bootstrap Filter

We have hidden states $CAR_t$, $R_t$, $I_t$, observed data $W_t$, $C_t$, and fixed parameter vector $\theta$. We are interested in learning the joint posterior distribution $P(CAR_{1:T}, R_{1:T}, I_{1:T}, \theta | W_{1:T}, C_{1:T})$. The goal of algorithm 1 (below) is to construct a set of particles $\left\{ \left( CAR_k^{(i)}, R_k^{(i)}, I_k^{(i)} \right) : i = 1, ..., N \right\}$ that approximate this distribution.

For simplicity, let our hidden states be collectively denoted $X_t = (CAR_t, R_t, I_t)$ and our observed data $y_t = (W_t, C_t)$. We start with the filtering distribution at time $t$ which we denote $q_t$ (defined below). We decompose this filtering distribution into the following recursion:

$$
\begin{aligned}
q_t &= P_\theta(X_{1:t}|y_{1:t}) \\
&\propto P_\theta(y_t|X_{1:t}, y_{1:t-1}) P_\theta(X_{1:t}|y_{1:t-1}) \\
&= P_\theta(y_t|X_{1:t}) P_\theta(X_t|X_{1:t-1}, y_{1:t-1}) P_\theta(X_{1:t-1}|y_{1:t-1}) \\
&= P_\theta(y_t|X_{1:t}) P_\theta(X_t|X_{1:t-1}) q_{t-1}
\end{aligned}
$$

where $P_\theta(y_t|X_{1:t}) = P_\theta(W_t|I_{1:t-1}) P_\theta(C_t|CAR_t, I_{1:t-1})$ is our joint observation distribution and $P_\theta(X_t|X_{1:t-1}) = P_\theta(CAR_t|CAR_{t-1}) P_\theta(R_t|R_{t-1}) P_\theta(I_t|I_{1:t-1})$ is our joint state-space transition distribution.

The decomposition makes two assumptions: (1) $y_t$ is conditionally independent of $y_{1:t-1}$ given $X_t$, and (2) $X_t$ is conditionally independent of $y_{1:t-1}$ given $X_{1:t-1}$. The definition of the observation and state-space transition distributions require further assumptions that are clear from their definition.

We define our importance sampling distribution $\pi$ in a similar recursive fashion:

$$
\pi(X_{1:t}|y_{1:t}) = \pi(X_t|X_{1:t-1}, y_{1:t}) \pi(X_{1:t-1}|y_{1:t-1})
$$

requiring the non-restrictive assumption that our importance distribution on $X_{1:t-1}$ does not depend on $y_t$. This further allows us to define the importance sampling weights recursively:

$$
\begin{aligned}
w_k^{(i)} &\propto \frac{P_\theta(y_t|X_{1:t}^{(i)}) P_\theta(X_t^{(i)}|X_{1:t-1}^{(i)}) q_{t-1}^{(i)}}{\pi(X_t^{(i)}|X_{1:t-1}^{(i)}, y_{1:t}) \pi(X_{1:t-1}^{(i)}|y_{1:t-1})} \\
&= \frac{P_\theta(y_t|X_{1:t}^{(i)}) P_\theta(X_t^{(i)}|X_{1:t-1}^{(i)})}{\pi(X_t^{(i)}|X_{1:t-1}^{(i)}, y_{1:t})} w_{k-1}^{(i)}
\end{aligned}
$$

Finally, we choose our importance sampling distribution $\pi(X_t^{(i)}|X_{1:t-1}^{(i)}, y_{1:t})$ to be the state-space transition distribution $P_\theta(X_t^{(i)}|X_{1:t-1}^{(i)})$, allowing these terms to be cancelled, and giving the recursion for the particle weights:

$$
w_k^{(i)} = P_\theta(y_t|X_{1:t}^{(i)}) w_{k-1}^{(i)}
$$

At each time step, we re-sample with replacement from our current particles according to $w_{k-1}^{(i)}$, thus the selected particles at time step $t$ can be viewed as direct draws from $P_\theta(X_t|y_{1:t})$.

Additionally re-sampling the $h$ most recent time steps re-weights past particles according to $P_\theta(y_t|X_{1:t}^{(i)})$, thus they can be viewed as samples from $P_\theta(X_t|y_{1:t+L})$. We note that in doing this resampling, we break the particle ancestry, so only have samples from the marginal distributions $P_\theta(X_t|y_{1:t+L}) \approx P_\theta(X_t|y_{1:T})$ and not the full joint distribution over state-trajectories $P_\theta(X_{1:t}|y_{1:t+L})$ (except for the final $L$ states).

Conditional on a fixed value of $\theta$, algorithm 1 presents our fixed-lag bootstrap filter. This assumes a default wind-in period equal to $h$ (30-days), although this can be changed as necessary.

#### 2.2 Practical Considerations for the Bootstrap Filter

##### 2.2.1 Choosing the Fixed-Lag $h$

In an ideal world we would resample the entire state-history at every step, producing direct samples from our desired posterior distribution $P(X_t|y_{1:T})$. In fact, this also admits samples from the joint posterior distribution across all time steps $P(X_{1:T}|y_{1:T})$, which would allow us to sample individual trajectories. However, as $T$ increases, the number of individual unique particles that remain in the earlier time steps (most obviously at $t = 1$) gets increasingly small. Eventually only a handful, or even a single, unique particle will remain for $X_1$, which provides a very poor approximation of $P(X_1|y_{1:T})$. This can be somewhat overcome by increasing the number of particles $N_x$, but given present computing power this quickly becomes impractical. Instead we re-sample only the most recent $h$ time steps, which means our particles at time step $t$ are samples from $P(X_t|y_{1:t+h})$ whenever $t + h < T$. The value of $h$ needs to be large enough that $P(X_t|y_{1:t+h}) \approx P(X_t|y_{1:T})$ while being small enough that we avoid particle degeneracy for reasonable values of $N_x$.

##### 2.2.2 Calculating Cumulative Infections

In figure 4 of the main text we present estimates of cumulative infections $CI_t$. If we were resampling entire particle histories at each time step (rather than fixed-lag resampling) we would be able to set $CI = \sum_{u=1}^{t} I_u^{(i)}$ for our $i^{th}$ sample of cumulative infections at time $t$. However, our fixed-lag resampling breaks the state-histories, so this approach is invalid. Instead, we augment cumulative infections as an additional hidden state, at each time step setting:

$$CI_t^{(i)} = CI_{t-1}^{(i)} + I_t^{(i)}$$

and resampling as usual. As the particle filter produces samples from $P(CI_t|y_{1:t+h})$, this method produces valid estimates of the pointwise cumulative infections $CI_t$.

**Algorithm 1** Fixed-lag bootstrap filter

Input: Parameter vector $\theta$, data $W_{1:T}$ and $C_{1:T}$
Sample $CAR_{-h:0}^{(i)}$, $R_{-h:0}^{(i)}$, and $I_{-h:0}^{(i)}$ from some initial state distribution
**for** $t = 1, ..., T$ **do**
  Sample $CAR_t^{(i)} \sim P_\theta(CAR_t | CAR_{t-1}^{(i)})$
  Sample $R_t^{(i)} \sim P_\theta(R_t | R_{t-1}^{(i)})$
  Sample $I_t^{(i)} \sim P_\theta(I_t | R_t^{(i)}, I_{t-L:t}^{(i)})$
  Set $w_t^{(i)} = P_\theta(W_t | I_{t-L:t}^{(i)}, CAR_t^{(i)}) P_\theta(C_t | CAR_t^{(i)}, I_{t-L:t}^{(i)})$
  Sample with replacement indices $\{x_j\}_{j=1}^N$ from $i = 1, 2, ..., N$ with probability $w_t^{(i)}$
  Update $(CAR_{t-L:t}^{(j)}, R_{t-L:t}^{(j)}, I_{t-L:t}^{(j)}) \leftarrow (CAR_{t-L:t}^{(x_j)}, R_{t-L:t}^{(x_j)}, I_{t-L:t}^{(x_j)})$
**end for**
Return $(CAR_{1:T}^{(i)}, R_{1:T}^{(i)}, I_{1:T}^{(i)})$

##### 2.2.3 Wind-In Period

In practice we wind-in using two steps: (1) hidden states are randomly allocated values for $t = -h$ to $t = 0$, and (2) the filter is run for $t = 1$ to $t = k$ for some $k$. We present results and calculate likelihoods for $t \geq k + 1$. The first step is necessary for there to be sufficient state-history to calculate the expected values of $I_t$, $C_t$, and $W_t$, which all involve convolutions of past infections. However, only a few of these randomly allocated trajectories will be plausible, leading to considerable uncertainty in the initial estimates of our hidden states. Thus we use data to filter particles as described above in the period $0 \leq t \leq k$ and start the estimation window at $t = k$, so that all particle chains have plausible past trajectories at this time. In general $k$ should be chosen to be greater than $h$.

This second wind-in period means that the estimation window only begins $k$ days after the start of the period for which data are available. It is possible to run the algorithm without the second wind-in period, which may be necessary when data are limited. However, this leads to greater uncertainty about estimated states in the early part of the time period and introduces substantial additional variation in the estimates of the model log-likelihood (section 2.3). In practice, we used $k = 50$, except for model runs starting on 1 January 2022 where we use $k = 31$ as the earliest available data were 1 December 2021.

#### 2.3 Likelihood Estimation

Thus far we have focused on estimating the value of the hidden states given some known parameter vector $\theta$. Our particle filtering algorithm also admits a tidy, albeit noisy, method for estimating the likelihood function $L(\theta|y_{1:T}) \propto P(y_{1:T}|\theta)$ [5]. First note that:

$$P(y_{1:T}|\theta) = \prod_{t=1}^{T} P_\theta(y_t|y_{1:t-1})$$

We can write each term in the product as:

$$P_\theta(y_t|y_{1:t-1}) = \int P_\theta(y_t|X_{t-L:t}) P_\theta(X_{t-L:t}|y_{1:t-1}) \, dX_{t-L:t}$$

Note $P_\theta(y_t|X_{t-L:t}^{(i}) = w_t^{(i)}$. Furthermore, our projected (but non-filtered) particles $\{\tilde{X}_{t-L:t}^{(i)}\}_{i=1}^N$ at time $t$ provide an approximation to $P_\theta(X_{t-L:t}|y_{1:t-1})$. Together this conveniently allows us to approximate this integral using:

$$P_\theta(y_t|y_{1:t-1}) \approx \frac{1}{N}\sum_{i=1}^{N} w_t^{(i)}$$

Taking logarithms gives us our estimator of the log-likelihood:

$$\hat{\ell}(\theta|y_{1:T}) = \sum_{t=1}^{T} \log\left(\frac{1}{N}\sum_{i=1}^{N} w_t^{(i)}\right)$$

As each term inside the outer sum is an approximation, the noise of this estimator grows with the length of data. In general this noise increases linearly with time, as does the time it takes to run a single filter, thus the computational requirements approximately scale with $O(T^2)$ [6].

##### 2.4 Particle Marginal Metropolis-Hastings

Particle marginal Metropolis-Hastings (PMMH; algorithm 2) is an established algorithm designed to estimate the joint posterior distribution $P(X_{1:T}, \theta|y_{1:T})$ of the hidden states and fixed parameter vector given our data, although in practice we use this method to estimate the marginal posterior distribution $P(\theta|y_{1:T})$. The algorithm uses the following proposal density:

$$q((X'_{1:T}, \theta')|(X_{1:T}, \theta)) = q(\theta'|\theta)P_{\theta'}(X'_{1:T}|y_{1:T})$$

where $X'_{1:T}$ are generated by running a particle filter at $\theta'$. This gives an acceptance probability of:

$$a = \min\left(1, \frac{\hat{P}(y_{1:T}|\theta')P(\theta')q(\theta|\theta')}{\hat{P}(y_{1:T}|\theta)P(\theta)q(\theta'|\theta)}\right)$$

where $q(\theta'|\theta)$ is our proposal density on our parameters and $\hat{P}(y_{1:T}|\theta)$ is an estimate of the model evidence at parameter vector $\theta'$ - this is the exponential of $\hat{\ell}(\theta|y_{1:T})$ described in section 2.3. The validity of using an estimate of the likelihood rather than an exact calculation is confirmed in [5], which is a key difference between PMMH and standard Metropolis-Hastings.

##### 2.5 Prior Distributions and Proposal Variances

The parameters $\sigma_R$, $\sigma_{CAR}$, $k_c$, and $k_w$ may change as the epidemiological landscape changes so we fitted them in five distinct time periods: (1) 1 January 2022 – 31 March 2022, (2) 1 April 2022 – 30 June 2022, (3) 1 July 2022 – 30 September 2022, (4) 1 October 2022 – 31 December 2022, and (5) 1 January 2023 – 31 March 2023. Choosing the duration of these windows requires balancing changing epidemiological dynamics (we expect these parameters to somewhat change over time) with using more data to obtain more precise estimates.

We use wide independent uniform distributions for our prior distributions on $\sigma_R$, $k_c$, and $k_w$. A wide uniform prior distribution can also be placed on $\sigma_{CAR}$, however, this results in a relatively

**Algorithm 2** Particle Marginal Metropolis-Hastings

**Require:** Prior distribution $P(\theta)$ on $\theta$, proposal density $q(\theta'|\theta)$, and number of MCMC steps $N$

Initialise $\theta_0 \sim P(\theta)$ and run algorithm 1 to estimate $\hat{P}_{\theta_0} = P(\theta_0|y_{1:T})$

**for** $i = 1, ..., N$ **do**

Sample $\theta' \sim q(\cdot|\theta_{i-1})$

Run algorithm 1 to estimate $\hat{P}_{\theta'} = P(\theta'|y_{1:T})$

Calculate acceptance probability $a_i = \min\left(1, \frac{\hat{P}(y_{1:T}|\theta')P(\theta')q(\theta_{i-1}|\theta')}{\hat{P}(y_{1:T}|\theta_{i-1})P(\theta_{i-1})q(\theta'|\theta_{i-1})}\right)$

Let $\theta_i = \theta'$ with probability $a_i$, else let $\theta_i = \theta_{i-1}$

**end for**

Return $(CAR^{(i)}_{1:T}, R^{(i)}_{1:T}, I^{(i)}_{1:T})$

high-valued posterior estimate for this parameter as the model can choose values of $CAR_t$ that closely fit the fluctuations in reported case data. We want our estimates of $CAR_t$ to reflect an underlying reporting rate, rather than the daily noise in reporting, so use a prior distribution on $\sigma_{CAR}$ to ensure this. For time periods encompassing 1 April 2022 – 31 March 2023 we use a normal distribution with mean 0.006 and standard deviation 0.00204, truncated on $(0, \infty)$, which has a $95^{th}$ quantile of 0.01. For the first time period, encompassing 1 January 2022 to 31 March 2022, we use a higher-mean normal distribution with mean 0.024 and standard deviation 0.00816, truncated on $(0, \infty)$, which has a $95^{th}$ quantile of 0.04. The use of a higher-mean prior distribution for $\sigma_{CAR}$ in this first period allows the model to fit to the rapid change in $CAR_t$ that is thought to have occurred when RATs were rolled out in February 2022 [4]. Table S1 reports our choices for prior distributions.

We use independent normal proposal densities for each parameter. The chosen standard deviations of the proposal densities are given in table S2. We outline how we chose these in section 2.6.

Table S1: Prior distributions on parameters. All normal distributions (represented by $N$) are truncated on $(0, \infty)$. The continuous uniform distribution is represented by $U$. The period starting 1 March 2022 continues until the end of considered period on 31 March 2023.

| Period starting | $\sigma_R$ | $\sigma_{CAR}$ | $k_c$ | $k_w$ |
|---|---|---|---|---|
| 1 Jan 2022 | N(0.024, 0.00816) | U(0, 0.1) | U(0, 400) | U(0, 0.02) |
| 1 Mar 2022 | N(0.006, 0.00204) | U(0, 0.1) | U(0, 400) | U(0, 0.02) |

Table S2: The chosen standard deviation for each independent normal proposal distribution.

| Period starting | $\sigma_R$ | $\sigma_{CAR}$ | $k_c$ | $k_w(\times 10^{-7})$ |
|---|---|---|---|---|
| 1 Jan 2022 | 0.024 | 0.004 | 4.3 | 1.4 |
| 1 Apr 2022 | 0.018 | 0.0014 | 21 | 5.5 |
| 1 Jul 2022 | 0.010 | 0.0015 | 30 | 6.6 |
| 1 Oct 2022 | 0.0073 | 0.001 | 22 | 7.4 |
| 1 Jan 2023 | 0.0089 | 0.0015 | 22 | 8.9 |

#### 2.6 Practical Considerations for Particle Marginal Metropolis-Hastings

In situations where the proposed particle is rejected, it is not necessary to re-estimate $\hat{P}_{\theta_{i-1}}$. One can typically let $\hat{P}_{\theta_i} = \hat{P}_{\theta_{i-1}}$. In some situations, particularly when the variance of the likelihood estimator is large, the Markov chain can get stuck on values of $\theta'$ where the estimate of $P_{\theta'}$ was unusually high, resulting in slower convergence. To avoid this we re-estimate $P_{\theta_{i-1}}$ if $n$ consecutive proposals have been rejected. Choosing $n$ requires balancing the computational cost of running additional particle filters with the cost of slower mixing chains. We use $n = 5$ in this work.

Theoretically, this algorithm works irrespective of the number of particles $N_x$ used in each filter, however, this does have a significant impact on the performance of the algorithm. A general heuristic is that $N_x$ should be chosen such that the standard deviation of $\hat{\ell}(\theta|y_{1:T})$ is around 1.2-1.3 [6]. This standard deviation is a function of $\theta$ itself (generally speaking estimates of $\ell$ at more likely values of $\theta$ have lower standard deviations) so choosing the ideal $N_x$ is not a simple task. We use $N_x = 10^5$ particles per filter when fitting to three-month time periods.

PMMH is a computationally expensive algorithm. Our results rely on 8 PMMH chains for each of the five time periods considered. Whilst running this on high-performance computing services can allow us to utilise the 40 cores required to run each chain simultaneously, it can still take multiple days to generate sufficient samples. We find that, with $N_x = 10^5$, it takes us approximately 20 hours to generate 2000 samples (although this can be faster if more modern central processing units are used). As seen in section 4, this can be enough to meet certain convergence criteria even if this is fewer samples than most MCMC algorithms target. Practically we expect this to make little difference to our posterior estimates of $\theta$, and even less of a difference to our posterior estimates of $X_{1:T}$. This final point can be seen by noting that the marginal posterior estimates of $X_{1:T}$ are very similar to the posterior estimates of $X_{1:T}$ conditional on any plausible value of $\theta$ - the majority of the uncertainty comes from the relationship between the hidden states and the observed data, rather than the hyper-parameters that characterise this relationship.

Despite generally using wide uniform prior distributions, we want to ensure our chains start at plausible values of $\theta$, otherwise considerable computation time must be spent on a wind-in period. Therefore, we first computed approximate bivariate heatmaps of the estimated log-likelihood on a coarse parameter mesh (see Supplementary Material sec. 3.2). We then initialised chains in the part of parameter space with relatively high likelihood values. As well as reducing convergence times, this technique also provides some reassurance that the PMMH algorithm is not missing any hidden modes, and that the posterior distribution found is similar to empirical results.

We ran the PMMH algorithm twice for each three-month block. First we did a training run, with seemingly plausible values for the parameter proposal variances, to provide a crude estimate of the posterior variance. Then a second, final, run was performed using the heuristic proposal variance of $2.38\sigma_{posterior}/n_{dim}$, where $\sigma_{posterior}$ is the estimated posterior standard deviation of the chosen parameter from the first run, and $n_{dim} = 4$ is the dimensionality of the parameter space. This heuristic could be replaced with adaptive MCMC methods - these are well known but slightly more complicated to implement from scratch.

#### 2.7 Posterior Distribution on Hidden States

One way of estimating the marginal posterior distribution $P(X_{1:T}|y_{1:T})$ is to store one (or more) trajectories from each PMMH step. The resulting set of particle trajectories are samples from the pointwise marginal posterior distribution $P(X_t|Y_{1:T})$. However, as we fit the parameters in three-month windows, the PMMH method only outputs trajectories in three-month blocks, which cannot be easily joined together.

Instead we first run algorithm 2 to generate a set of fixed parameter values $\{\theta_i\}_{i=1}^{N_c} \sim P(\theta|y_{1:T})$. We then sample from $P(X_{1:T}|y_{1:T})$ by iteratively uniformly sampling $\theta^*$ from $\{\theta_i\}_{i=1}^{N_c}$, running algorithm 1 with $N_x$ particles at $\theta = \theta^*$, and keeping $N_s$ trajectories (where $N_s \leq N_x$) from the output. Repeating this $N_c$ times (once for each parameter sample) gives a set of $N_s N_c$ particle trajectories that approximate the pointwise posterior distribution $P(X_t|y_{1:t+h}) \approx P(X_t|y_{1:T})$. Note that each sample from $\{\theta_i\}_{i=1}^{N_c}$ consists of five independent sets of values for the inferred parameters ($\sigma_{CAR}$, $\sigma_R$, $k_c$ and $k_w$), one for each of the five three-month periods. When we run algorithm (2) for the whole 15-month period, we assume that the values of these four parameters change instantaneously from one three-month block to the next.

Typically we want to run this with sufficient unique draws from $P(\theta|y_{1:T})$ (say $N_c \geq 100$) to appropriately account for uncertainty in $\theta$. The number of samples retained from each iteration should be chosen so our overall number of trajectories is sufficiently large - we choose $N_c N_s = 2 \times 10^6$. Finally, as we are less concerned by minor degeneracy in individual particle filters, the number of particles used in each filter $N_x$ can be smaller than if we were just running the filter once. For our results, $N_x = 10^5$ worked well, but this needs to be tailored to individual purposes.

When presenting results we calculate the mean of the samples as the central estimate and use the $2.5^{th}$ and $97.5^{th}$ quantiles to represent our 95% credible intervals (CrI).

#### 2.8 Pre-Determined Parameters

In addition to the estimated parameters, there are others that we fix. These are the generation time distribution $g_u$, infection-to-reporting distribution $L_u$, the infection-to-shedding distribution $\omega_u$, and the average total genome copies per infection $\alpha$. We also pre-specify the fixed-lag resampling window $h = 30$.

The generation time is assumed to be a discretised Gamma random variable with mean 3.3 days and standard deviation 1.3 days [4, 7–9]. The infection-to-reporting and infection-to-shedding distributions are calculated as convolutions of an incubation period (infection-to-onset) distribution (Weibull with mean 2.9 days and standard deviation 2.0 days [10]) and an onset-to-reporting distribution (estimated from New Zealand data, mean 1.8 days and standard deviation 1.8 days) or an onset-to-shedding distribution (mean 0.7 and standard deviation 2.6 [1]). The Gamma and Weibull distributions were discretised by taking their value at integer times and normalising. All of these distributions are presented in Supplementary Figure S1.

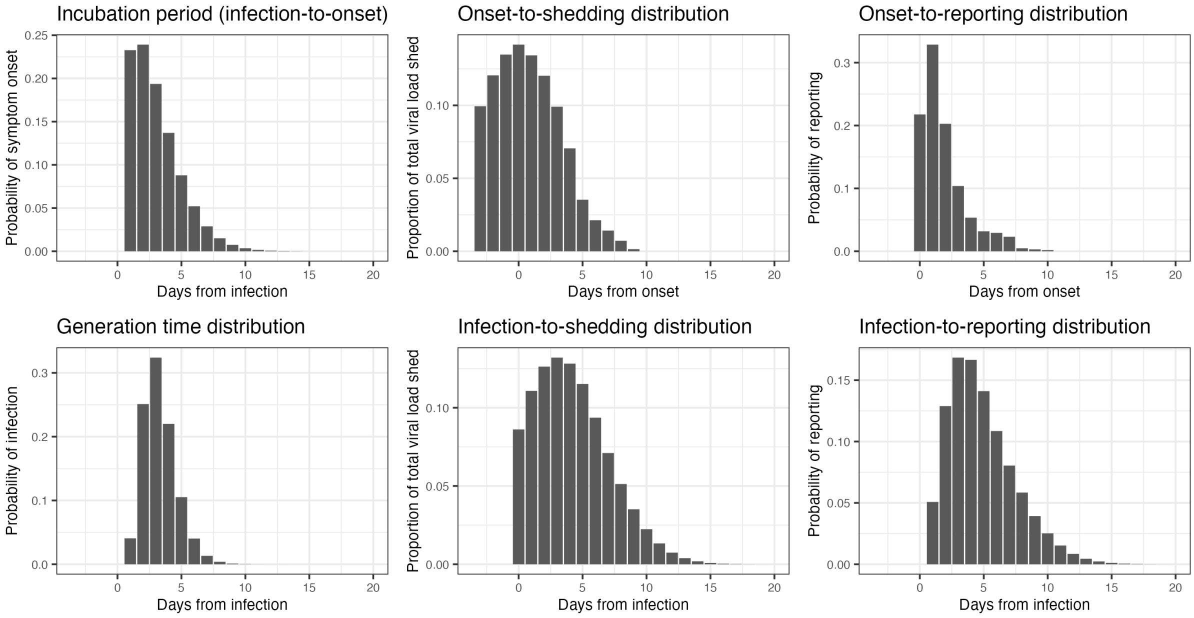

Figure S1: Delay distributions. Distributions reported in the first row were used as inputs to create the infection-to-shedding and infection-to-reporting distributions reported in the second row.

##### 2.9 Estimating Curvewise Extrema

Our primary methods produce pointwise estimates of the hidden states at each time step (e.g. mean and quantiles of the samples at each fixed value of $t$). As the timing of peaks and troughs can be quite variable between particles, using pointwise statistics to quantify the heights and timings of peaks or troughs (e.g. local maxima in the median value across particles) can be misleading [11]. For this purpose, is important to consider these on a curvewise basis (e.g. median of the local maxima across particles). This requires samples from the joint posterior distribution $P(X_{s:t}|y_{1:T})$ over some fixed window $s : t$.

We achieve this by increasing our resampling length $h$ and halting the algorithm when the period of interest is contained in $(T-h, T-30)$ where $T$ represents the final stopping time. Limiting the lower window to $t \geq T-h$ ensures that complete trajectories are faithfully resampled over the time period of interest. Limiting the upper window to $t \leq T-30$ ensures we are using appropriately smoothed samples that are sufficiently informed by data.

### 3 Supplementary Results

##### 3.1 Synthetic Verification of Hidden State Estimates

Before analysing real-world data, we performed synthetic tests to verify our model. We imposed a prescribed time-varying reproduction number and CAR and ran a forward simulation of our model to calculate the median number of infections, cases and wastewater data for a fixed value

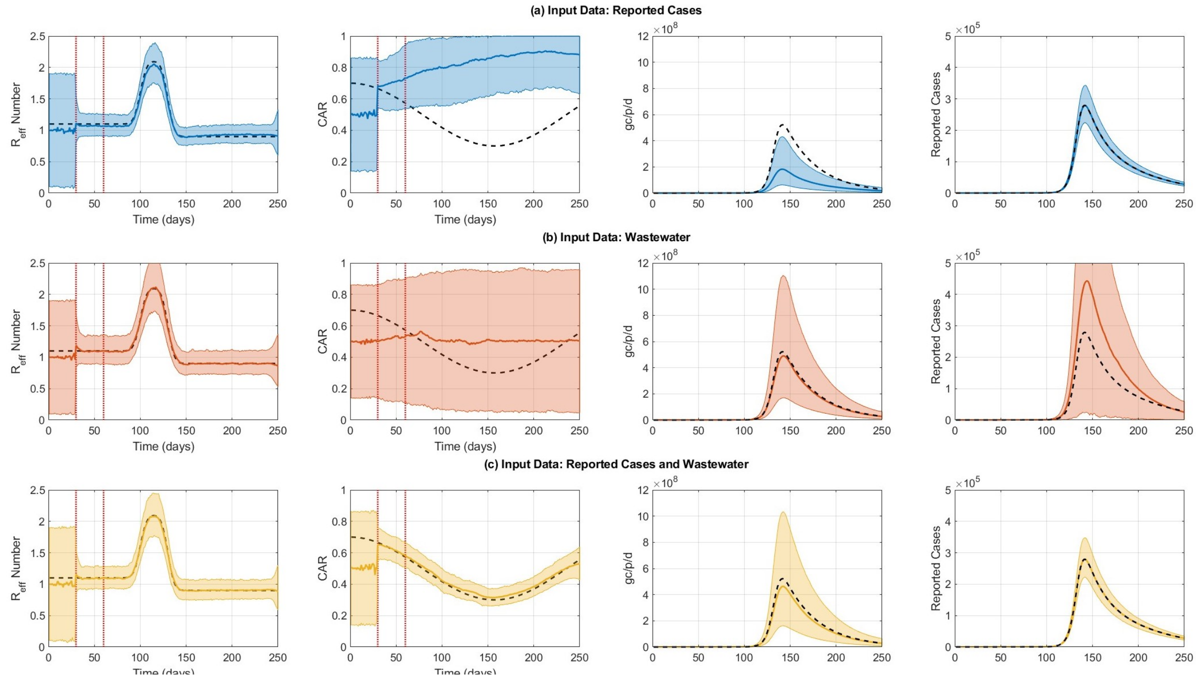

Figure S2: Synthetic results showing (left) instantaneous reproduction number, (centre-left) case ascertainment rate, (centre-right) wastewater data in genome copies per person per day (gc/p/d) and (right) reported cases. Results are shown when using (a) only reported cases as input data for the particle filter, (b) only wastewater data, and (c) both reported cases and wastewater data. Solid lines present central estimates. Shaded regions show 95% CrIs on the value of the hidden states (left and centre-left columns) and 95% CrIs on the prediction distribution for wastewater data and reported cases (centre-right and right columns). Black dashed lines indicate the synthetic data. Vertical red lines in hidden state plots (left and centre-left columns) indicate the end of the two wind-in periods (see Supplementary Materials sec. 2.2).

of $\alpha$. We then used the simulated case and wastewater data as inputs to the particle filter to estimate $R_t$ and $CAR_t$. To investigate how the addition of wastewater data affected model performance, we ran the particle filter with three different sets of inputs: only case data, only wastewater data, and both case and wastewater data. The results are shown in Figure S2. The parameters used were $\sigma_R = 0.1$, $\sigma_{CAR} = 0.02$, $k_c = 100$, and $k_w = 1 \times 10^{-6}$ along with $\alpha = 3 \times 10^9$.

All three data combinations resulted in a reasonable estimate of $R_t$ (Figure S2). The model error was smallest when using both reported cases and wastewater data as input (root mean square error between the true solution and the median of the particle filter output was 3.4 and 0.7 times larger when only using reported cases or wastewater data, respectively). Previous work has estimated $R_t$ from reported case data [12]. The results presented in Figure S2 demonstrate that $R_t$ can also be estimated from wastewater data independently from case data and that the most accurate result is achieved by combining reported case information with wastewater data.

For CAR, there were significant differences between the three different sets of input data (Figure S2). When using only reported cases or wastewater data separately, the model did not have sufficient information to inform estimates of CAR. As a consequence, estimates were either inaccurate or did not capture the temporal trend and had very wide credible intervals. When reported case information was combined with wastewater data, there was good agreement between the estimated CAR and the true solution, with a relatively narrow credible interval. This illustrates the value of combining wastewater data with reported case information to obtain reliable estimates of changes in CAR over time.

##### 3.2 Visualising Log-Likelihood Estimates

As discussed in section 2.6, we simulated the model likelihood on a coarse grid of parameter values to get a preliminary estimate of the plausible range of parameter values. We present examples of two outputs in figure S3. The left-plot shows log-likelihood estimates for $\sigma_R$ for the third estimation window (1 July 2022 to 30 September 2022) while the right-plot shows a bivariate heatmap of log-likelihood estimate for $\sigma_{CAR}$ and $k_c$. Code to reproduce these figures is provided in the *usefulscripts* subfolder at `https://github.com/nicsteyn2/NZWastewaterModelling`.

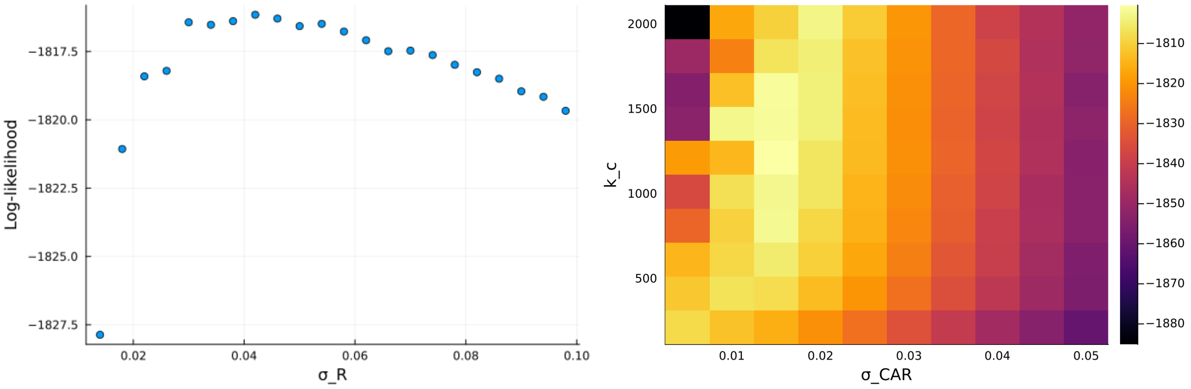

Figure S3: Log-likelihood estimates for various values of $\sigma_R$ (left) and various combinations of values of $\sigma_{CAR}$ and $k_c$ (right).

##### 3.3 Sensitivity to Delay Distributions

The model relies upon three distributions describing the delay from infection to reporting, shedding detected viral genome copies, and infecting other people. We test the effect of shifting these distributions by one day backward or forward. We do this by appending a zero at the start (to shift times backward), or by removing the first entry in $d$ (to shift times forward), where $d_i$ is the probability vector specifying the likelihood the delay takes $i$ days. This has the effect of shifting the mean of the distributions by approximately one day in either direction.

Figures S4 to S6 show the effect of these shifts to be minimal, with the exception of the shedding distribution, where shifts can substantially impact estimates of the absolute case ascertainment rate, even though the relative case ascertainment rate is similar despite the shifts.

Note, due to computational limitations we do not re-fit fixed parameters for each shift, although the effect of doing this is expected to be negligible.

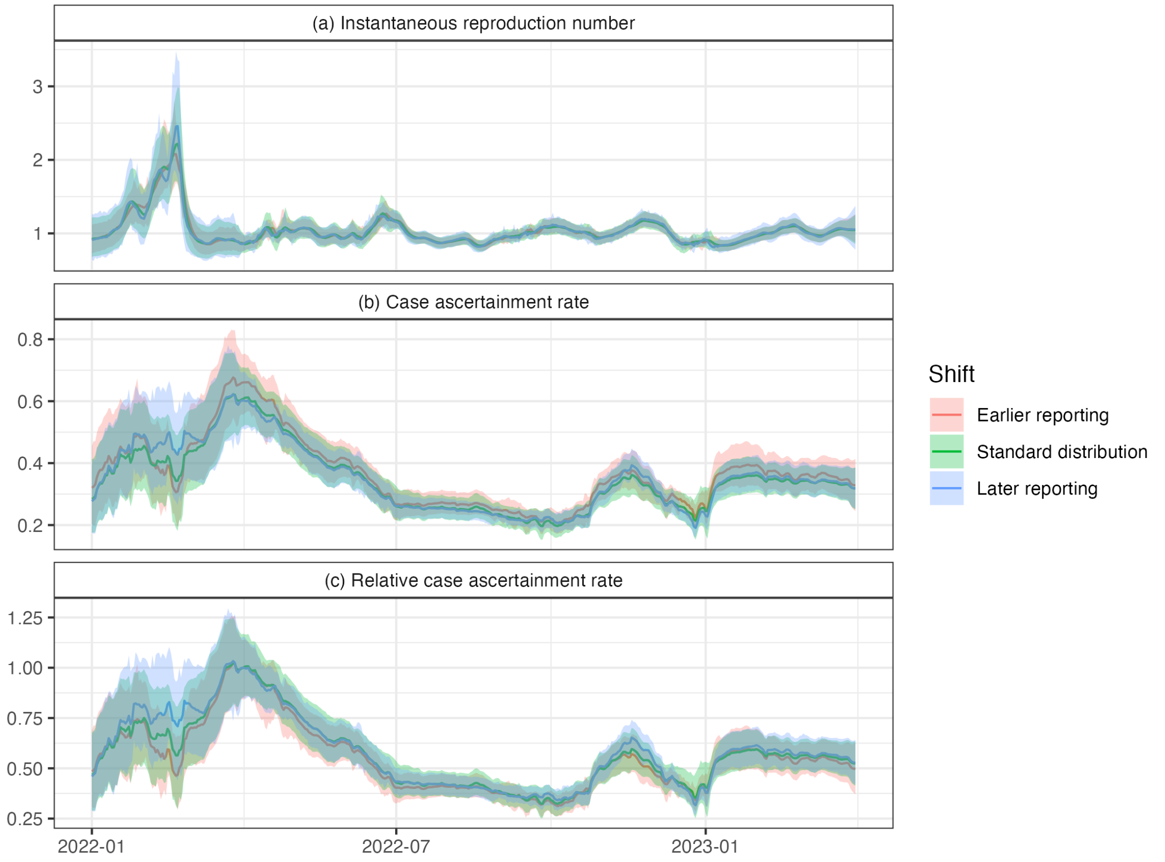

Figure S4: The effect of shifting the reporting time distribution by one day forward or backward.

##### 3.4 Fitting to reported cases and wastewater data separately

The observation distribution is the product of the case observation distribution and the wastewater observation distribution. We can instead run the model with only one of these observation distributions. As there is no longer any information about the case ascertainment rate, we fix this at an arbitrary value of $CAR_t = 0.5$ with no fluctuation over time ($\sigma_{CAR} = 0$). We use PMMH to again fit the relevant parameters to each model (cases-only and wastewater-only). We report the posterior mean and 95% credible intervals, alongside estimates from the full model, in Table S3.

In general, estimates of $\sigma_R$ were greater when fit to a single source of data. For reported cases, this was accompanied by slightly smaller estimates of $k_c$, suggesting that the assumption of a fixed $CAR$ caused the attribution of some uncertainty to shift from the observation process to fluctuations in the hidden state $R_t$.

Figure S7 shows reproduction number estimates from these separate models. The credible intervals resulting from fitting the model to wastewater data alone are substantially wider than when fitting to cases and wastewater. The reproduction number estimates from fitting the model to cases alone exhibit higher short-term fluctuations at times, which could be suggestive of overfitting.

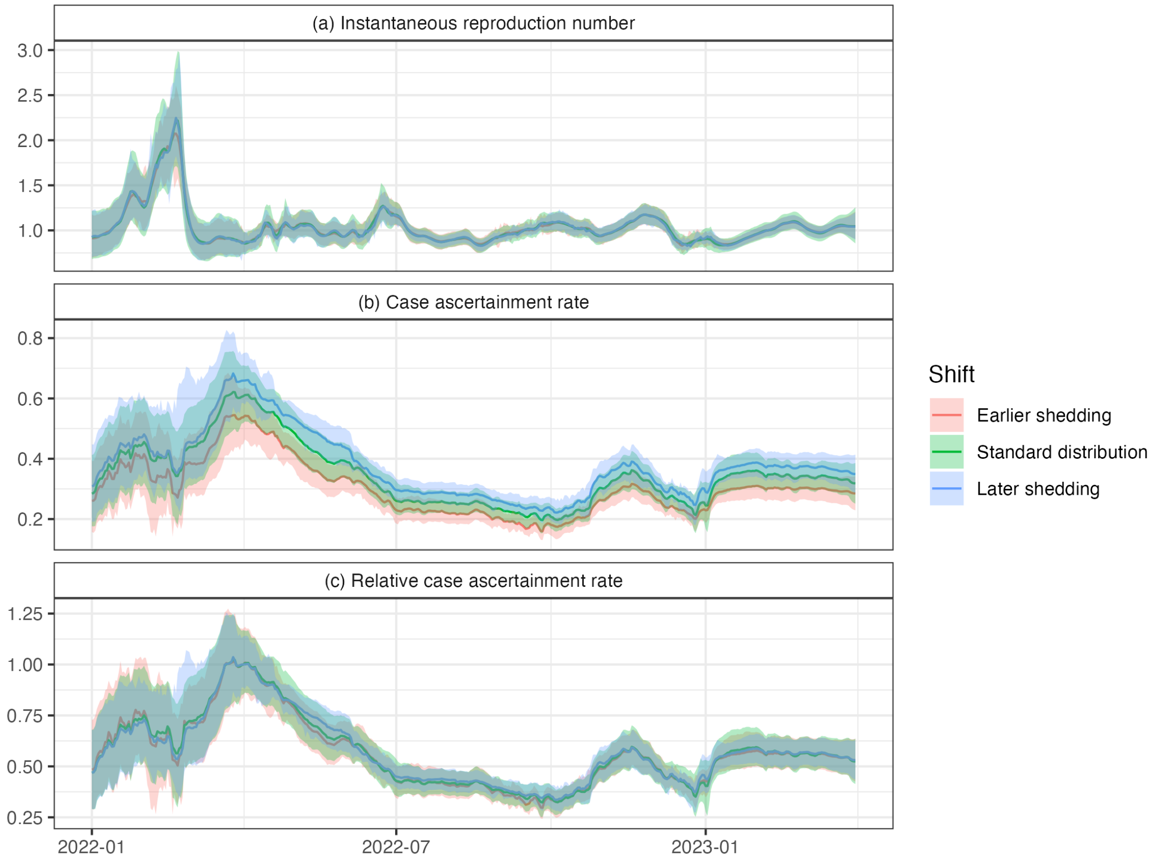

Figure S5: The effect of shifting the shedding time distribution by one day forward or backward.

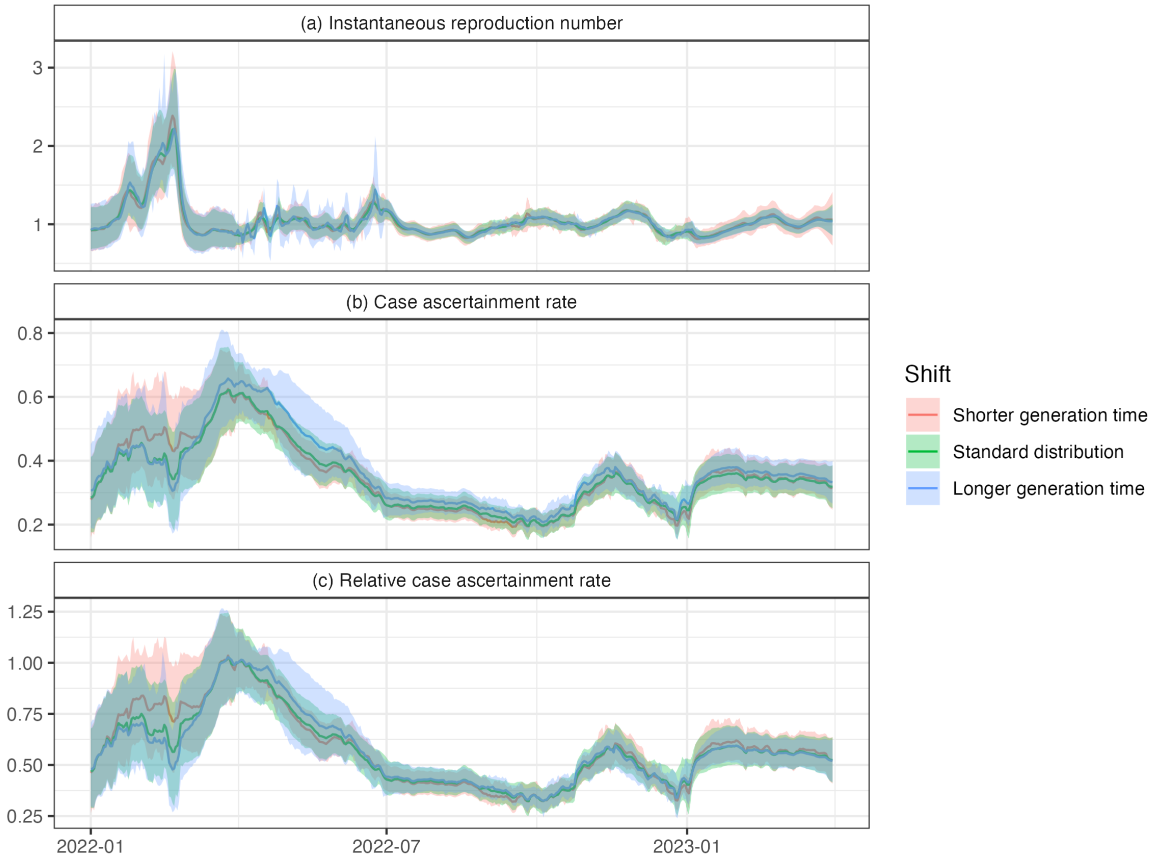

Figure S6: The effect of shifting the generation time distribution by one day forward or backward.

Table S3: Central estimates and 95% CrIs for estimated model parameters in each time period for the model when fit to both series, as well as separately to cases and wastewater data. Dates in the 'Period' column are the start date for the three-month period. All outputs presented to 2 s.f. Higher values of $\sigma_R$ and $\sigma_{CAR}$ suggest $R_t$ and $CAR_t$ vary faster. Higher values of $k_c$ and $k_w$ indicate a lower variance in the corresponding observation distribution. Note a different prior distribution was used for $\sigma_{CAR}$ in the first period (see Supplementary Material, sec. 2.5), which may also impact estimates of other parameters in this period.

| Period | Model | $\sigma_R$ | $\sigma_{CAR}$ | $k_c$ | $k_w(\times 10^{-6})$ |
|---|---|---|---|---|---|
| 1 Jan 2022 | Joint | 0.12 (0.069, 0.21) | 0.03 (0.017, 0.043) | 31 (20, 49) | 1.5 (1.1, 2) |
| | Cases | 0.14 (0.077, 0.23) | 0 (assumed) | 27 (18, 38) | - |
| | Wastewater | 0.14 (0.070, 0.25) | 0 (assumed) | - | 1.5 (1.1, 2.1) |
| 1 Apr 2022 | Joint | 0.069 (0.041, 0.12) | 0.0099 (0.0053, 0.014) | 170 (100, 250) | 4.8 (3.2, 6.8) |
| | Cases | 0.056 (0.030, 0.11) | 0 (assumed) | 150 (100, 210) | - |
| | Wastewater | 0.057 (0.036, 0.10) | 0 (assumed) | - | 5.2 (3.6, 7.3) |
| 1 Jul 2022 | Joint | 0.037 (0.020, 0.066) | 0.0063 (0.0018, 0.01) | 330 (220, 400) | 4.8 (3.3, 6.5) |
| | Cases | 0.053 (0.022, 0.13) | 0 (assumed) | 240 (140, 360) | - |
| | Wastewater | 0.060 (0.024, 0.14) | 0 (assumed) | - | 5.2 (3.7, 7.1) |
| 1 Oct 2022 | Joint | 0.038 (0.020, 0.068) | 0.011 (0.0073, 0.014) | 170 (110, 270) | 7.2 (4.7, 10) |
| | Cases | 0.048 (0.020, 0.098) | 0 (assumed) | 120 (64, 140) | - |
| | Wastewater | 0.063 (0.032, 0.12) | 0 (assumed) | - | 8.5 (5.7, 12) |
| 1 Jan 2023 | Joint | 0.038 (0.018, 0.073) | 0.0093 (0.0041, 0.015) | 150 (84, 330) | 6.8 (4.4, 10) |
| | Cases | 0.056 (0.023, 0.11) | 0 (assumed) | 140 (71, 270) | - |
| | Wastewater | 0.059 (0.027, 0.12) | 0 (assumed) | - | 7.1 (4.9, 10) |

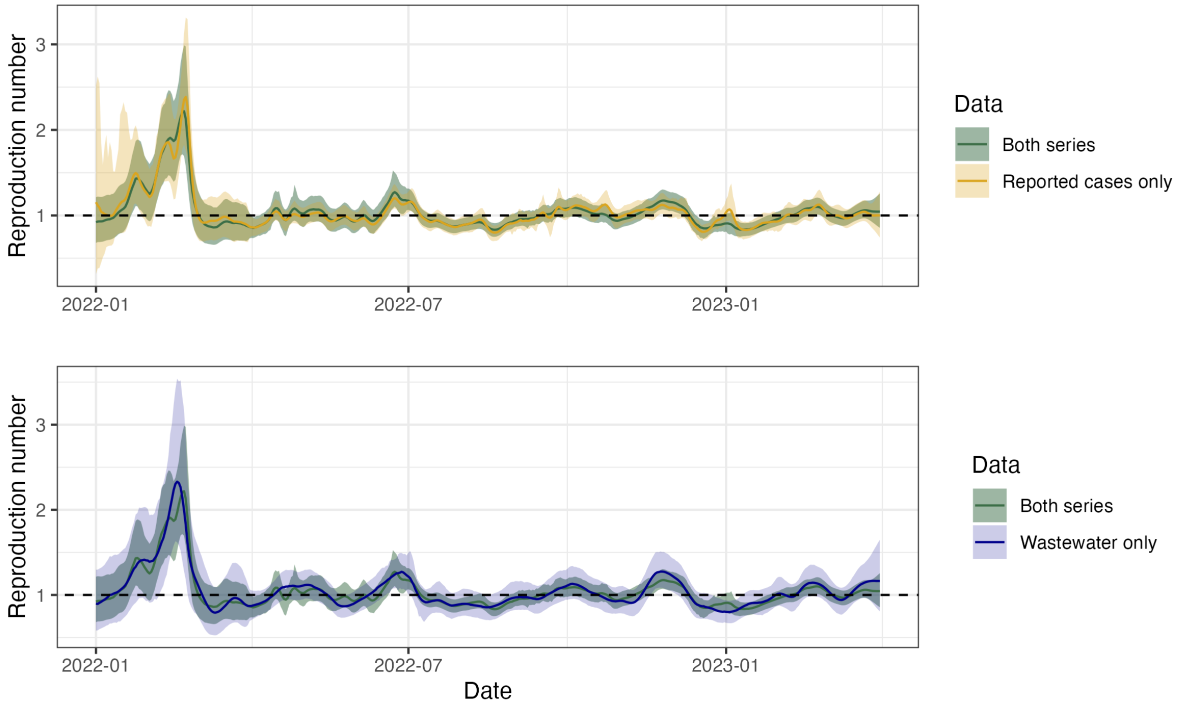

Figure S7: The estimated reproduction number when the model is fit to reported cases only (gold, upper panel) and wastewater data only (blue, lower panel). The original method fit to both series is included for comparison (green, both panels).

### 4 Particle Marginal Metropolis-Hastings Outputs

This section presents the trace plots, pairwise scatterplots of samples, pairwise correlations of samples, univariate kernel density plots, and the Gelman-Rubin diagnostic for each of the four parameters over the five three-month time periods in which they are estimated. Each of the eight chains were run for 5,000 iterations, with the initial 100 samples dropped as a wind-in period (although as discussed in section 2.6, our initial choices of parameter values mean this is somewhat superfluous).

The Gelman-Rubin diagnostic tests for convergence by comparing intra-chain variance with inter-chain variance. We use the *coda* package in R [13] to calculate a point estimate and 95% upper confidence bound on the Gelman-Rubin diagnostic. It is generally accepted that values less than 1.1 imply convergence, although some use the more relaxed cutoff of 1.2. These are reported in table S4.

Table S4: Gelman-Rubic diagnostics and total sample sizes (for all eight chains) for each parameter and time period.

| Period starting | $\sigma_R$ | $\sigma_{CAR}$ | $k_c$ | $k_w$ | Sample size |
|---|---|---|---|---|---|
| 1 Jan 2022 | 1.01 (1.01) | 1.01 (1.02) | 1.01 (1.01) | 1.00 (1.02) | 39,200 |
| 1 Apr 2022 | 1.00 (1.01) | 1.01 (1.06) | 1.01 (1.02) | 1.01 (1.02) | 39,200 |
| 1 Jul 2022 | 1.01 (1.02) | 1.01 (1.02) | 1.01 (1.03) | 1.01 (1.01) | 39,200 |
| 1 Oct 2022 | 1.01 (1.02) | 1.01 (1.04) | 1.02 (1.04) | 1.00 (1.01) | 39,200 |
| 1 Jan 2023 | 1.02 (1.04) | 1.02 (1.05) | 1.04 (1.09) | 1.01 (1.03) | 39,200 |

#### 4.1 Period 1: 1 January 2022 – 31 March 2022

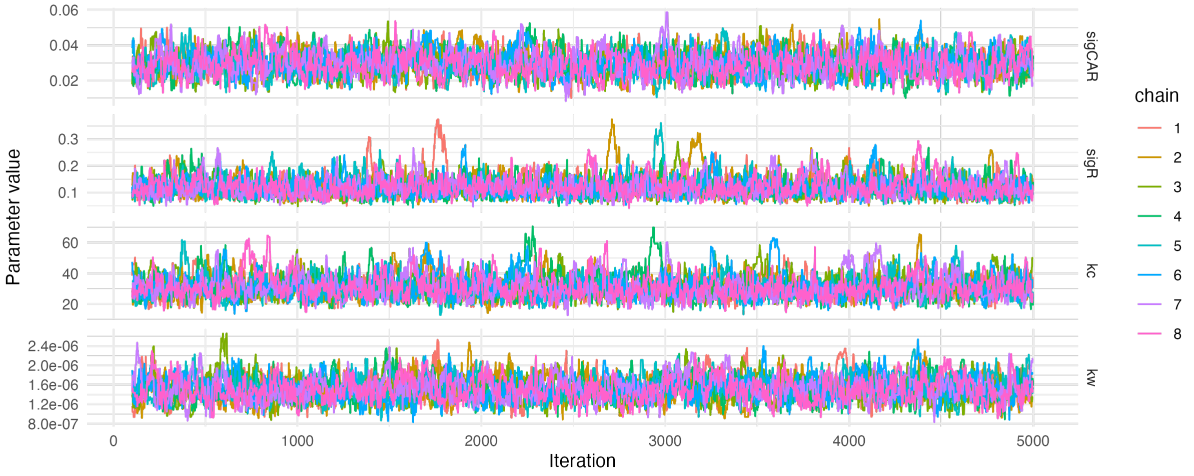

Figure S8: MCMC trace plots for period 1.

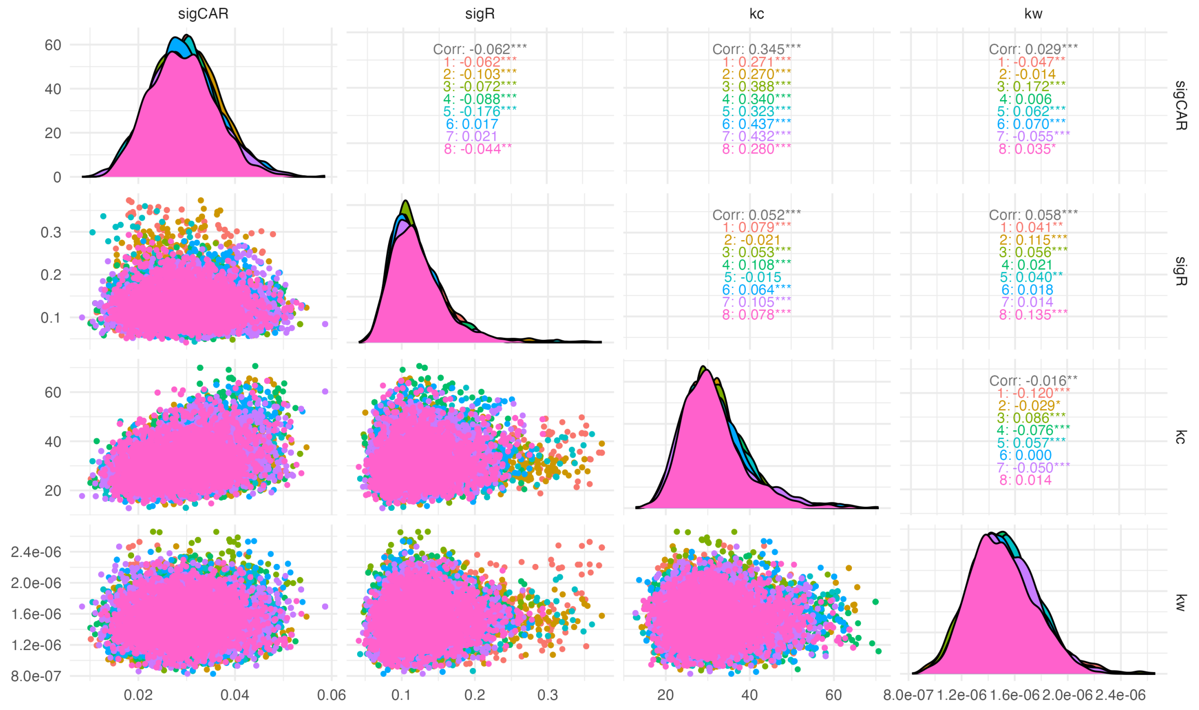

Figure S9: MCMC pairwise scatter plots, univariate kernel densities, and pairwise correlation estimates for period 1.

#### 4.2 Period 2: 1 April 2022 – 30 June 2022

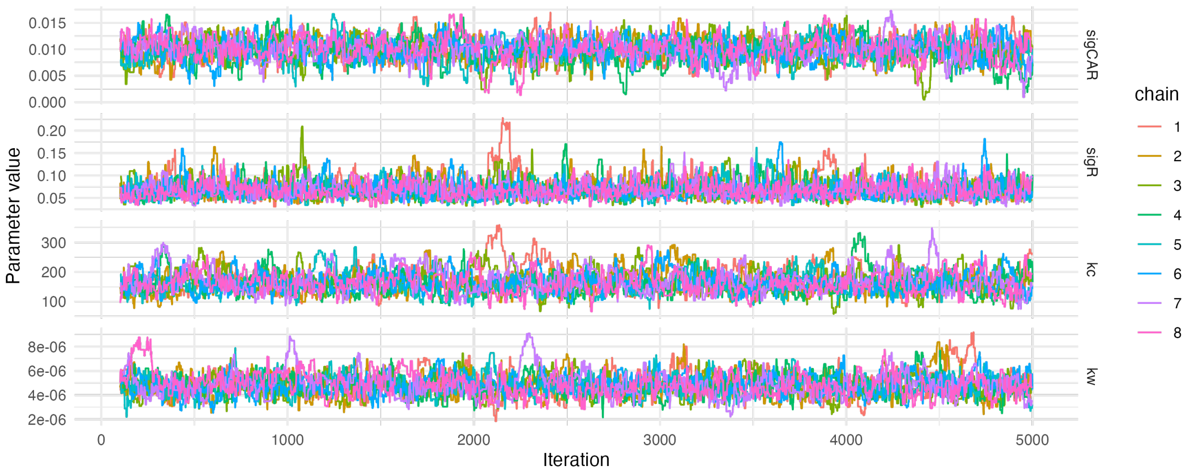

Figure S10: MCMC trace plots for period 2.

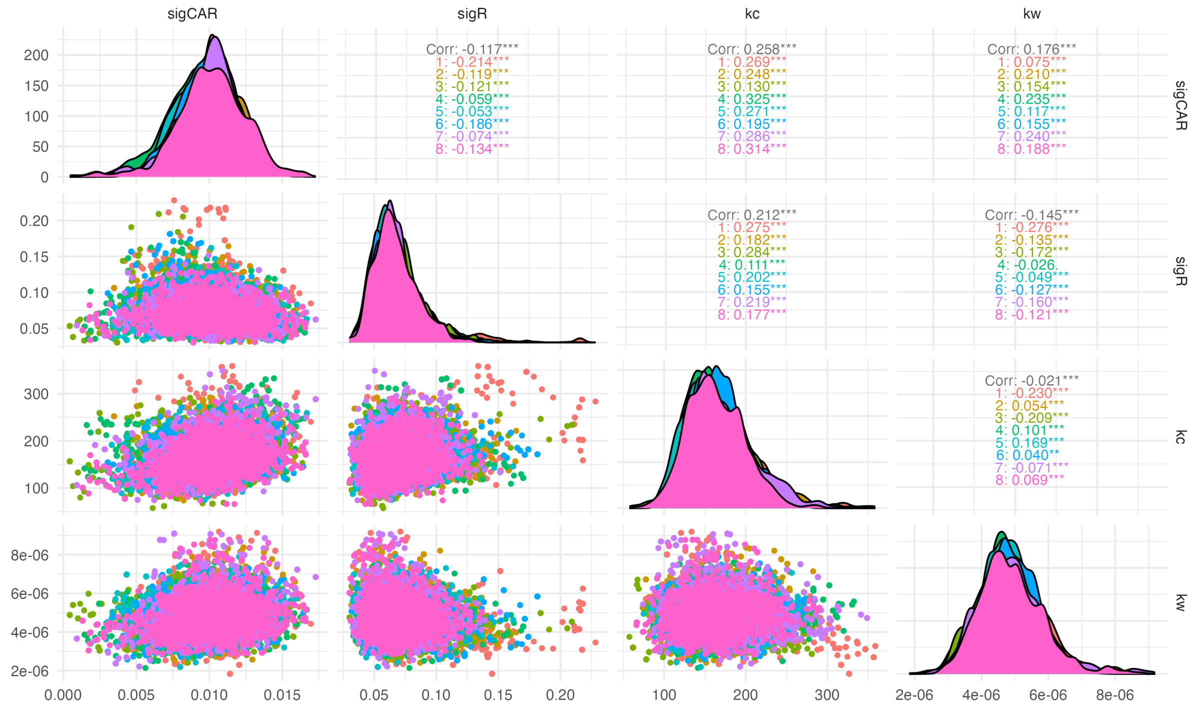

Figure S11: MCMC pairwise scatter plots, univariate kernel densities, and pairwise correlation estimates for period 2.

#### 4.3 Period 3: 1 July 2022 – 30 September 2022

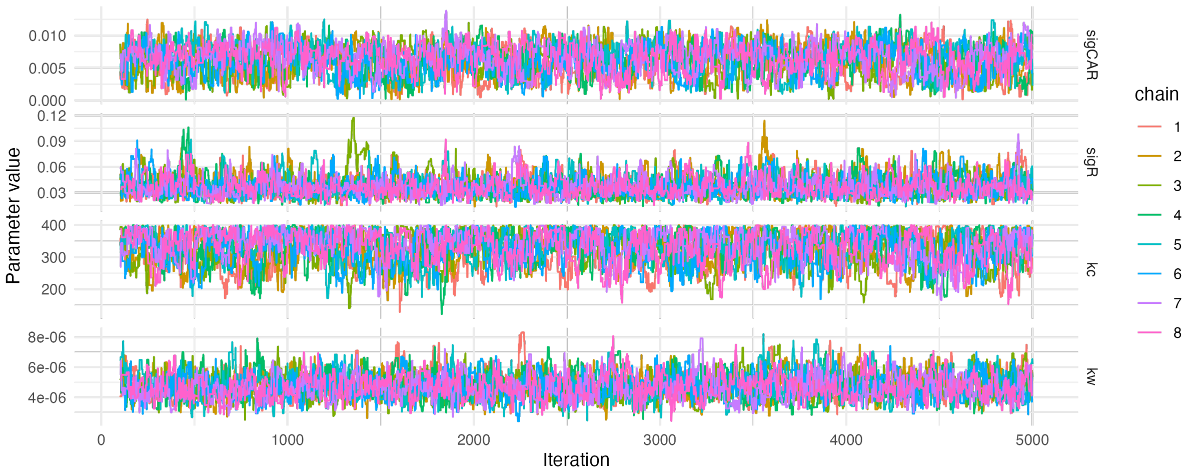

Figure S12: MCMC trace plots for period 3.

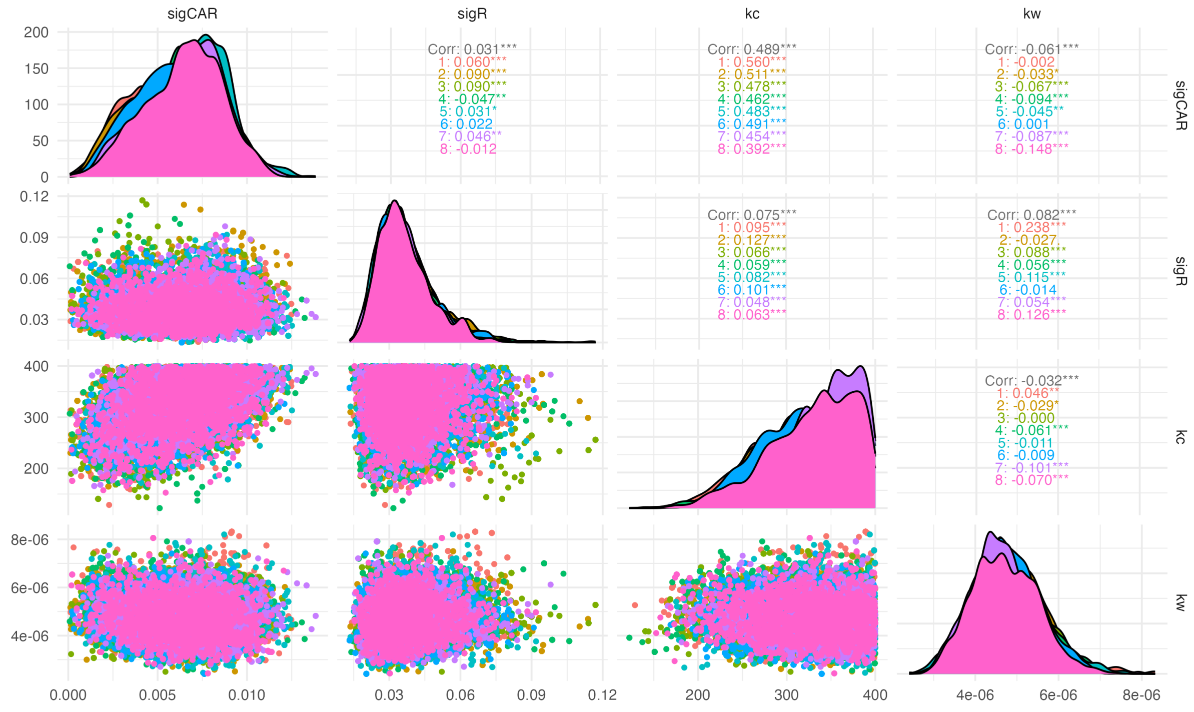

Figure S13: MCMC pairwise scatter plots, univariate kernel densities, and pairwise correlation estimates for period 3.

#### 4.4 Period 4: 1 October 2022 – 31 December 2022

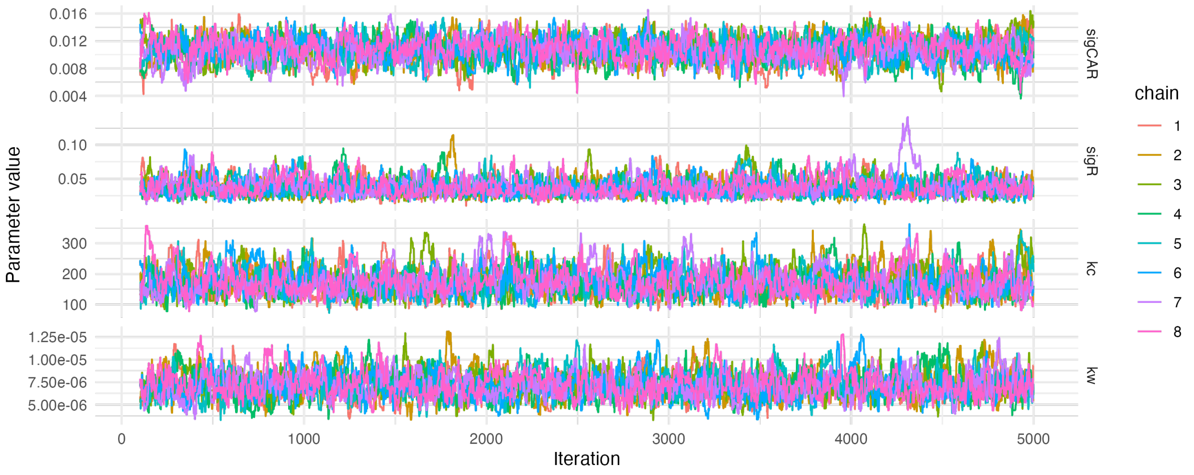

Figure S14: MCMC trace plots for period 4.

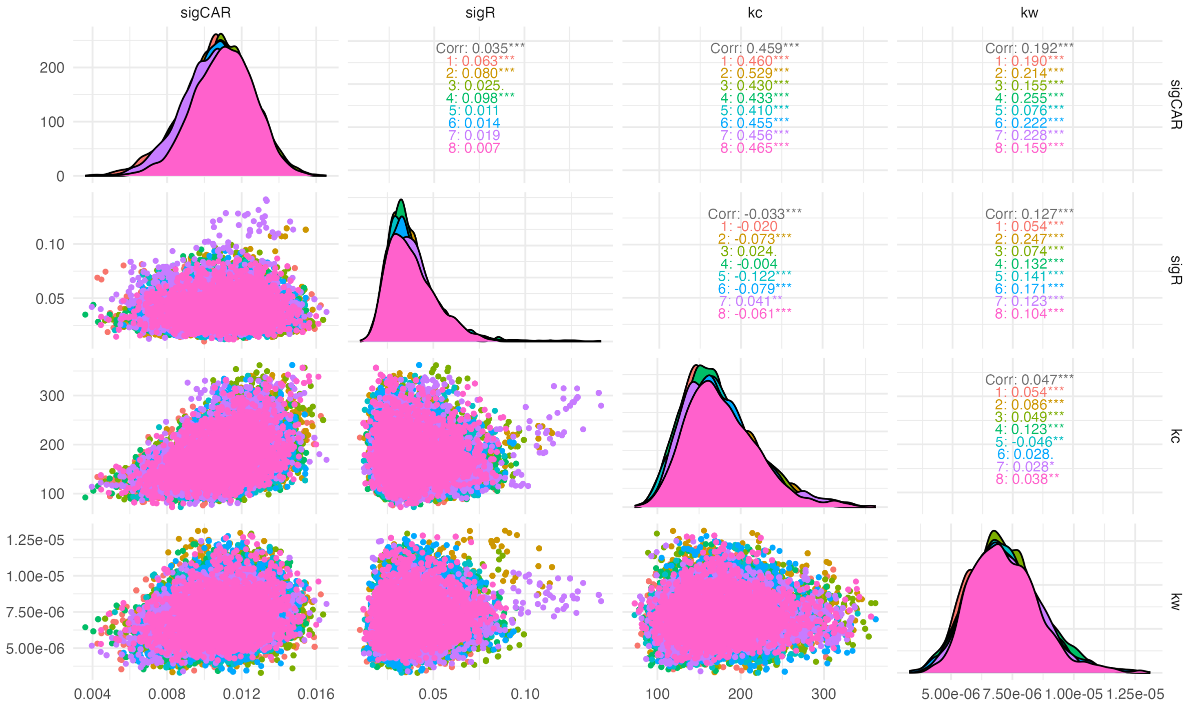

Figure S15: MCMC pairwise scatter plots, univariate kernel densities, and pairwise correlation estimates for period 4.

#### Period 5: 1 January 2023 - 31 March 2022

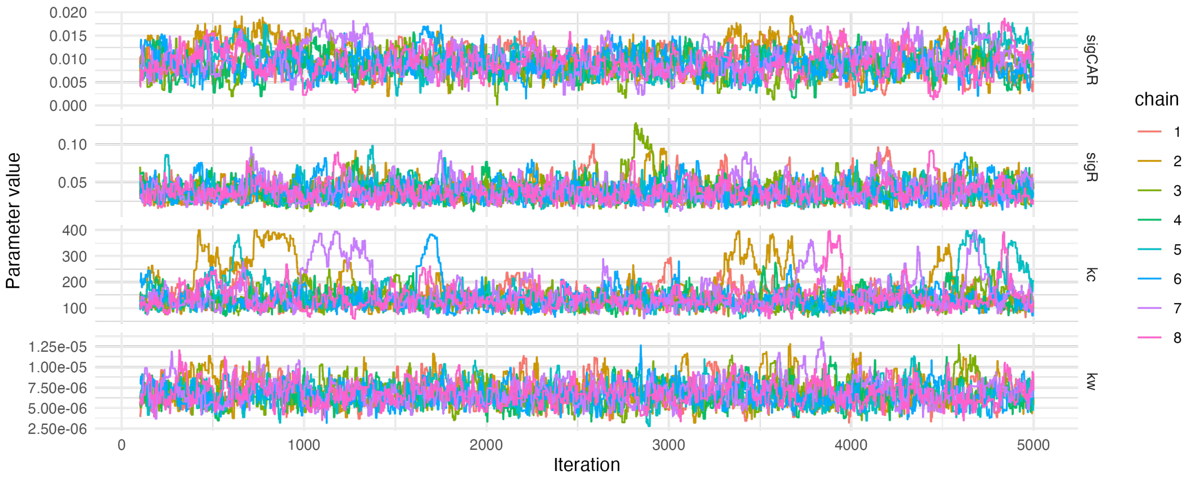

Figure S16: MCMC trace plots for period 5.

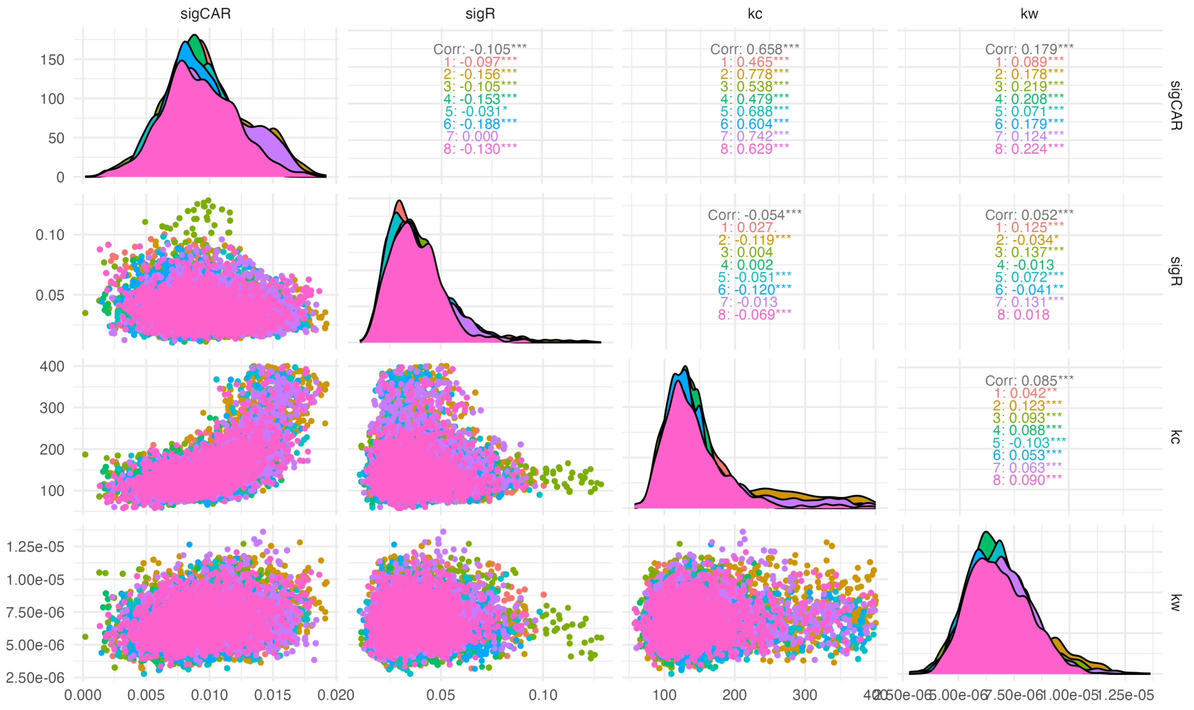

Figure S17: MCMC pairwise scatter plots, univariate kernel densities, and pairwise correlation estimates for period 5.
